# Treatment with human umbilical cord-derived mesenchymal stem cells for COVID-19 patients with lung damage: a randomised, double-blind, placebo-controlled phase 2 trial

**DOI:** 10.1101/2020.10.15.20213553

**Authors:** Lei Shi, Hai Huang, Xuechun Lu, Xiaoyan Yan, Xiaojing Jiang, Ruonan Xu, Siyu Wang, Chao Zhang, Xin Yuan, Zhe Xu, Lei Huang, Jun-Liang Fu, Yuanyuan Li, Yu Zhang, Weiqi Yao, Tianyi Liu, Jinwen Song, Liangliang Sun, Fan Yang, Xin Zhang, Bo Zhang, Ming Shi, Fanping Meng, Yanning Song, Yongpei Yu, Jiqiu Wen, Qi Li, Qing Mao, Markus Maeurer, Alimuddin Zumla, Chen Yao, Wei-Fen Xie, Fu-Sheng Wang

**Affiliations:** Department of Infectious Diseases, Fifth Medical Center of Chinese PLA General Hospital, National Clinical Research Center for Infectious Diseases, Beijing, China; Wuhan Huoshenshan Hospital, Wuhan, China; Department of Respiratory, Changzheng Hospital, Second Military Medical University, Shanghai, China; Optical Valley Branch of Maternal and Child Hospital of Hubei Province, Wuhan, China; Department of Hematology, Second Medical Center of Chinese PLA General Hospital, Beijing, China; Peking University Clinical Research Institute, Peking University First Hospital, Beijing, China; Department of Infectious Disease, General Hospital of Central Theater Command, Wuhan, China; VCANBIO Cell & Gene Engineering Corp., Ltd. Tianjin, China; National Industrial Base for Stem Cell Engineering Products, Tianjin, China; Department of Hematology, Union Hospital, Tongji Medical College, Huazhong University of Science and Technology, Wuhan, China; Key Laboratory of Cancer Center, Fifth Medical Center of Chinese PLA General Hospital, Beijing, China; Department of Endocrinology and Metabolism, Changzheng Hospital, Second Military Medical University, Shanghai, China; Department of Radiology, Union Hospital, Tongji Medical College, Huazhong University of Science and Technology, Wuhan, China; Nursing Department, Fifth Medical Center of Chinese PLA General Hospital, Beijing, China; Department of Gastroenterology, Changzheng Hospital, Second Military Medical University, Shanghai, China; Immunotherapy Programme, Champalimaud Centre for the Unknown, Lisbon, Portugal; I Med Clinic, University of Mainz, Mainz, Germany; Center for Clinical Microbiology, Division of Infection and Immunity, University College London, and UCL Hospitals NIHR Biomedical Research Centre, London, UK

## Abstract

**BACKGROUND:** Treatment of severe Corona Virus Disease 2019 (COVID-19) is challenging. We performed a phase 2 trial to assess the efficacy and safety of human umbilical cord-mesenchymal stem cells (UC-MSCs) to treat severe COVID-19 patients with lung damage, based on our phase 1 data.

**METHODS:** In this randomized, double-blind, and placebo-controlled trial, we recruited 101 severe COVID-19 patients with lung damage. They were randomly assigned to receive either UC-MSCs (4 × 10^7^ cells per infusion) or placebo on day 0, 3, and 6. The primary endpoint was an altered proportion of whole lung lesion volumes from baseline to day 28. Other imaging outcomes, 6-minute walk test, maximum vital capacity, diffusing capacity, and adverse events were recorded and analysed.

**RESULTS:** 100 COVID-19 patients were finally recruited to receive either UC-MSCs (n = 65) or placebo (n = 35). UC-MSCs administration exerted numerical improvement in whole lung lesion volume from baseline to day 28 compared with the placebo (the median difference was -13.31%, 95%CI -29.14%, 2.13%, P=0.080). UC-MSCs significantly reduced the proportions of solid component lesion volume compared with the placebo (median difference: -15.45%; 95% CI -30.82%, -0.39%; P=0.043). The 6-minute walk test showed an increased distance in patients treated with UC-MSCs (difference: 27.00 m; 95% CI 0.00, 57.00; P=0.057). The incidence of adverse events was similar in the two groups.

**CONCLUSIONS:** UC-MSCs treatment is a safe and potentially effective therapeutic approach for COVID-19 patients with lung damage. (Funded by The National Key R&D Program of China and others. ClinicalTrials.gov number, NCT04288102.)

## Introduction

The Coronavirus Disease 2019 (COVID-19), caused by severe acute respiratory syndrome coronavirus 2 (SARS-CoV-2) infection,^1^ causes substantial damage to lungs, ranging from mild respiratory illness to severe acute respiratory syndrome, and death,^2-4^ Dysregulated immune responses of both the innate and adaptive immune systems are associated with disease severity, lung damage and long term functional disability.^5-8^ There are currently no prophylactic vaccines or effective antiviral agents available to treat COVID-19 and management of COVID-19 patients remains largely symptomatic and supportive therapy.^9^ Therefore, there is an urgent need for safe and alternative therapeutic options to mitigate inflammatory organ injury. Currently ongoing clinical trials of immunotherapeutic approaches include convalescent plasma therapy, monoclonal antibodies against interleukin-6, and cellular therapies.^10-12^ Mesenchymal stem cells (MSC) are non-hematopoietic cells with immune modulatory, regenerative, and differentiation properties.^13^ MSC treatment reduced the pathological changes of the lung and inhibits the cell-mediated immune-inflammatory response induced by the influenza virus in animal models and clinical trials.^14 15^ The safety and potential efficacy of MSC have also been evaluated in the patients with acute respiratory distress syndrome (ARDS).^16-19^ The immunomodulatory and regenerative properties of MSCs offer potential cellular therapeutic option for limiting lung damage in patients with COVID-19 and require evaluation in randomized controlled. In a phase 1 trial we previously demonstrated that intravenous transfusions of human umbilical cord (UC)-MSCs in patients with moderate and severe COVID-19 were safe and well tolerated (NCT04252118).^20^ We now report results of a randomized, double-blind, placebo-controlled trial performed at 2 medical centers in Wuhan, China, evaluating the safety and efficacy of intravenous treatment with UC-MSCs in severe COVID-19 patients with lung damage.

## Methods

### Design

We conducted a randomized, placebo-controlled, double-blind phase 2 trial (ClinicalTrials.gov: NCT04288102). The study was done between 5 March 2020 and 28 March 2020. Ethical approval was obtained from the institutional review boards of each participating hospital. Written informed consent was obtained from all the enrolled patients or their legal representatives if they were unable to provide consent. The clinical protocol and statistical analysis plan are available in Supplement 1 and Supplement 2.

### Inclusion and Exclusion Criteria

Hospitalised patients with severe COVID-19 with laboratory-confirmed SARS-CoV-2 infection by reverse transcription polymerase chain reaction (RT-PCR) were screened. Patients were eligible if they met any of the following criteria:1) severe COVID-19 diagnosed after onset of disease; 2) chest computed tomography (CT) imaging confirmed pneumonia combined with lung damage. The illness severity of COVID-19 was evaluated in accordance with Guidelines issued by the National Health Commission of China (version 7.0).^21^ Briefly, patients with any of the following conditions but without invasive ventilation, shock or other organ failure (need organ support therapy) were considered as severe cases: 1) dyspnoea (respiratory rate ≥ 30 times/min); 2) oxygen saturation of 93% or lower on room air; 3) arterial oxygen partial pressure (PaO_2_)/fraction of inspired oxygen (FiO_2_) ≤ 300 mmHg; 4) pulmonary imaging showing that the foci progressed by > 50% in 24-48 hours. The exclusion criteria included patients with shock or COVID-19 combined with any one of other organ failures, those who received invasive ventilation, or patients with any malignant tumour, pregnancy or breastfeeding, or co-infection of other pathogens.

### Randomization and masking

Eligible patients were randomly assigned in a 2:1 ratio to receive either UC-MSCs or the placebo, in addition to standard care, using an interactive web response management system (IWRS). A permutated-block randomisation sequence that was stratified by the trial sites was generated and uploaded to the system. Patients, investigators, and outcome assessors (independent central imaging reviewers) were all blinded to the treatment allocation. Blinding was also ensured by the product marking, with the UC-MSCs and the placebo having a similar appearance and packaging.

A barcode-based product management system (Product Identification Authentication and Tracking System, PIATS) was introduced in this study to manage and track the study products logistics, e.g., preparation, packaging, shipping, storage, and clinical administration to the patients. The application of PIATS could realize the blind label processing by non-informative unique barcodes of the clinical study drug. The concealment of the randomisation sequence could be ensured using PIATS and IWRS.

### UC-MSCs preparation, dosage, and safety monitoring

UC-MSCs were prepared by VCANBIO Cell & Gene Engineering Corp, Tianjin, China. Briefly, the MSCs were obtained according to the method described in our previous study.^20^ The UC-MSCs used in this study came from one umbilical cord of full-term deliveries (after consultation with the parents of UC donor). The Wharton’s Jelly (WJ) tissues were cut into approximately 2 mm^3^ pieces from cord tissue and planted upside down on tissue culture flasks (75□cm^2^) cultured in DMEM/F12, supplemented with fetal bovine serum (10% FBS, BI, Israel) at 37°C with 5% CO_2_. The adherent cells were detached with 1×TrypLE (GIBCO, USA) and then re-plated at a density of approximately 6-8×10^3^ cells/cm^2^ for further expansion. Master cell bank at passage 2 and working cell bank at passage 4 were set separately. A homogenous population of cultured cells at passage 5 were prepared as UC-MSC product. The culture cells were identified by the minimal criteria suggested by International Society for Cellular Therapy(ISCT): (1) plastic adherent under tissue culture flask; (2) >95% of the cell population expressed CD105, CD73 and CD90, and these cells were lack expression (<2% positive) of CD45, CD34, CD11b, CD19 and HLA-DR as measured by flow cytometry (BD, FACS Calibur, USA); (3) differentiation potential into osteoblasts, adipocytes and chondroblasts under standard in vitro differentiating conditions. For each individual MSC batch, cell viability was examined using both trypan blue and 7-AAD/Annexin V staining by flow cytometry after preparation in Tianjin and before intravenous transfusion in Wuhan, respectively (Supplement 3). The cell product has been certified by the National Institutes for Food and Drug Control of China.

The UC-MSC product was an almost colourless suspension containing 4.0 × 10^7^ MSCs in a volume of 100 ml/bag. The placebo had the same medium and appearance in packaging and suspension, but without the MSCs. After preparation, both the MSC and placebo products were shipped to the clinical facilities in an ice box with a real-time monitoring and alarm device for temperature and location to ensure the best storage conditions (8–12 □). Shipping of cell products by express railway from Tianjin to Wuhan took less than 6 hours.

The treatment dose was 4.0×10^7^ cells for each procedure, and three procedures were carried out for each patient on day 0, 3, and 6 after randomisation. Infusion was started with a standard blood filter tubing set with a pore size of 170 μm. Under electrocardiographic monitoring, the cell product was infused by gravity within 60 min. We also monitored continuous pulse oximetry as well as patient’s physical signs including body temperature, pulse, skin color, respiration, and blood pressure during the infusion period and up to 30 min after infusion.

The incidence and nature of all adverse events were reviewed and assessed by the investigators to determine whether they were related to the administration of the study product. Methods for data collection and in-study measurements are described in detail in Supplement 1.

### Imaging and clinical outcomes

All patients underwent high-resolution chest CT examination at baseline, day 10, and day 28. The primary outcome was gauged as a change in the total lesion proportion (%) of the whole lung volume from baseline to day 28, as measured by chest CT. It was defined as (total lesion proportion of the whole lung volume at day 28–total lesion proportion of the whole lung volume at baseline) / total lesion proportion of the whole lung volume at baseline. The secondary imaging outcomes were a change in the total lesion proportion (%) of the whole lung volume from baseline to day 10, a change in solid component and ground-glass lesion proportion from baseline to day 10, 28, and change in lung densitometry at day 10, 28. Lung lesions were evaluated by using the changes in high-resolution chest CT images and measured by centralised imaging interpretation based on both lung radiologist analyses and imaging software. The imaging data were derived from a software-assisted lung volumetry and densitometry procedure (Supplement 1).

Clinical outcomes within 28 days included 6-minute walk test (6-MWT), status of oxygen therapy maximum forced vital capacity (VC_max_), diffusion lung capacity for carbon monoxide (DL_CO_), modified Medical Research Council Dyspnoea Scale (mMRC), changes in absolute lymphocyte counts and subsets, as well as plasma cytokine and chemokine levels. Safety evaluation included adverse events and all-cause mortality. Detailed definitions and assessment procedures are described in Supplement 1.

### Statistical analysis

This study was designed as a phase 2 clinical trial. The limited efficacy information of the medication in patients with COVID-19 and the exploratory nature of this study meant that the original target sample size was not justified by statistical calculation and was set as 45 patients, with an allocation ratio of 2:1. Minimal serious adverse events were observed in our phase 1 trial; therefore, the sample size was expanded to 90, and finalized at 101, to obtain more data from this study. Sample size adjustments were made in a manner that maintained the double-blind status of this study and were approved by the institutional review boards of the two participating hospitals.

There were no pre-defined hypotheses made in this study; therefore, we focused on description instead of inference for statistical analyses: all statistical tests, confidence intervals, and *P*-values were used for exploration, not for inference. For primary outcome analysis – the change in the total lesion proportion (%) of the whole lung volume from baseline to day 28 and the difference between the UC-MSC and placebo groups was tested using wilcoxon rank sum test and the median differences were calculated using the Hodges–Lehmann estimation (It was also applied to other secondary outcomes which were not in accordance with normal distribution.). Six category scale and MMRC dyspnea score were calculated by using ordinal logistic regression model. The modified intention-to-treat (mITT) population was considered as the primary analysis population and safety analysis was done in all patients who started their assigned treatment. If the patient missed a chest CT scan, the last scan’s results were carried forward to the missing visit for primary endpoints in the mITT analysis. Other missing values of secondary endpoints and per-protocol analyses were not imputed. Statistical analyses were performed using SAS software, version 9.4 (Cary, NC, USA). The figures were generated using GraphPad Prism 7 software (GraphPad Inc., La Jolla, CA, USA).

## Results

### Study Population

From March 5, 2020, to March 28, 2020, a total of 288 patients were screened at two hospitals in Wuhan city. The majority of severe hospitalized COVID-19 patients were at the convalescent stage and some of them were with progression stage. Among them, 101 eligible patients previously diagnosed as severe COVID-19 type, being referred as the ITT population, were randomized in a 2:1 ratio (66 to the UC-MSC group and 35 to the placebo group). One patient in the treatment group withdrew her previously written informed consent after randomisation and did not receive UC-MSC infusion. Therefore, 65 and 35 patients were treated with UC-MSC or placebo, respectively, who were defined as the modified intention-to treat (mITT) population, as shown in Figure 1. Some patients missed the follow-up check on day 28 or received the examination outside of the follow-up window; therefore, the per-protocol (PP) population included 49 patients in the MSC group and 25 patients in the placebo group (Supplement 4).

**Figure 1:**
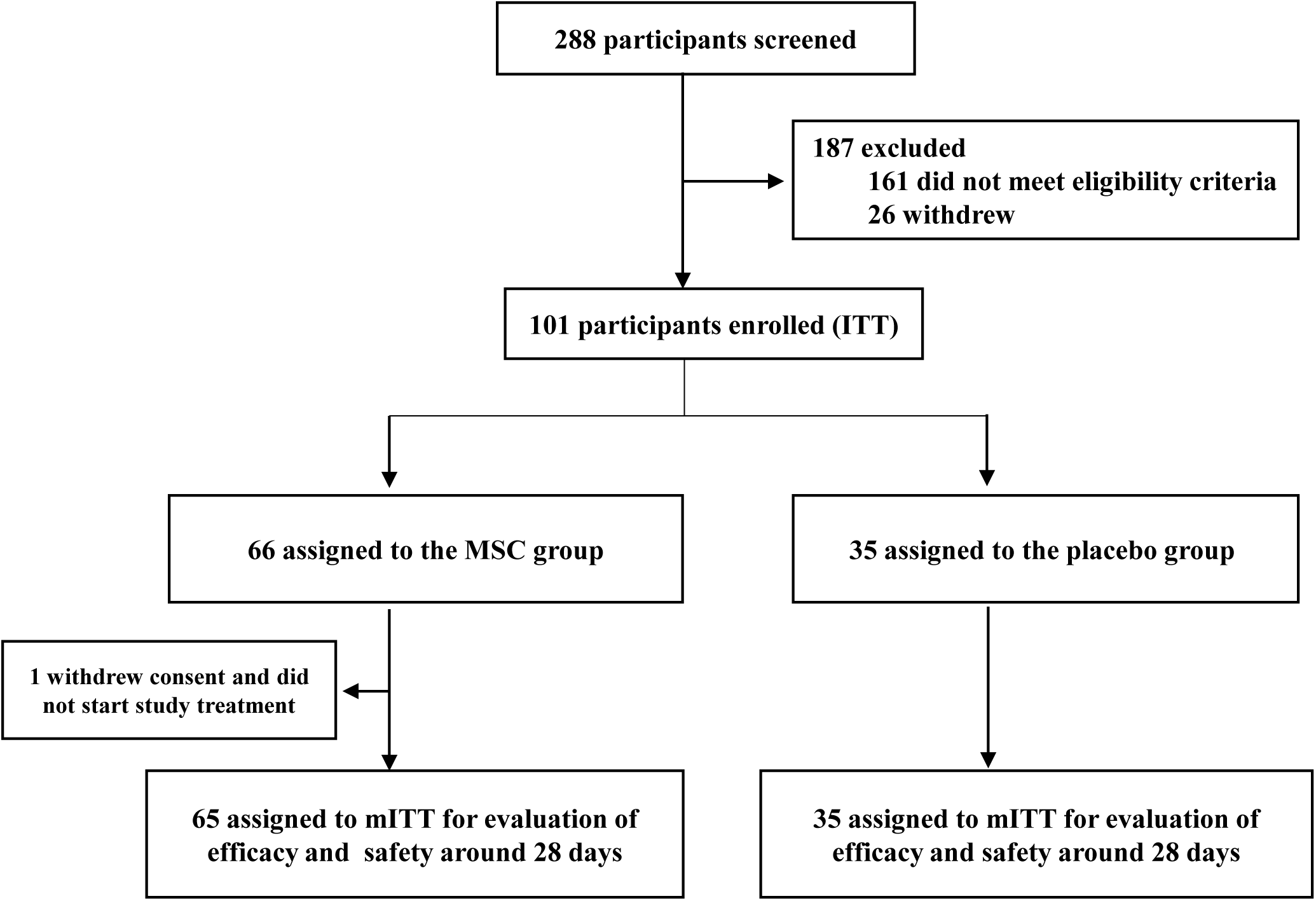
Trial profile. ITT= intention-to treat population. mITT= modified intention-to treat population

### Baseline characteristics

The baseline characteristics were highly consistent between the two groups of patients in the mITT population (Table 1). Briefly, baseline age, sex, BMI, time from symptom onset, distribution of comorbidities, concomitant medication, and lesion proportions assessment from chest CT were matched in the two groups. The median time from symptoms onset to study baseline was 45.00 (39.00, 51.00) days in the MSC group and 47.00 (41.00, 53.00) days in the placebo group. The most common comorbidity was hypertension, followed by diabetes. The majority of the patients at baseline has a clinical status of category 2 or category 3, as shown by evaluation of the Six Category Scale. In detail, there were 14 (21.54%), 50 (76.92%), and 1 (1.54%) patients in category 2, 3, and 4, respectively, at baseline in the MSC group; and 10 (28.57%), 25 (71.43%), 0 (0.00%) in category 2, 3, and 4, respectively, in the placebo group. There were no statistical differences in laboratory results, including D-dimer, interleukin-6 (IL-6), and C-reactive protein (CRP) between the two groups.

**Table 1:** Baseline patient characteristics.

|  | UC-MSc group (n = 65) | Placebo group (n = 35) |
| --- | --- | --- |
| Age, years | 60.72(9.14) | 59.94(7.79) |
| Sex – no. (%) |  |  |
| Men | 37 (56.92%) | 19 (54.29%) |
| Women | 28 (43.08%) | 16 (45.71%) |
| BMI (Body Mass Index), Kg/m<sup>2</sup>* | 24.71(3.19) | 25.01(3.12) |
| Time from symptom onset to baseline, days | 45.00(39.00,51.00) | 47.00(41.00,53.00) |
| Any comorbidities | 34 (52.31%) | 18 (51.43%) |
| Hypertension | 17 (26.15%) | 10 (28.57%) |
| Diabetes | 12 (18.46%) | 5 (14.29%) |
| Chronic bronchitis | 2(3.08%) | 3(8.57%) |
| Chronic obstructive pulmonary disease | 2(3.08%) | 0(0.00%) |
| Concomitant medication |  |  |
| Antiviral drugs | 32 (49.23%) | 20 (57.14%) |
| Antibiotics | 27 (41.54%) | 12 (34.29%) |
| Corticosteroids | 13 (20.00%) | 9 (25.71%) |
| Lesion proportion (%): total lesion volume (in cm<sup>3</sup>) / whole lung volume (in cm<sup>3</sup>) | 26.31(11.62,38.42) | 27.98(11.57,44.14) |
| Solid component lesion proportion (%): Solid component lesion volume (in cm<sup>3</sup>) / whole lung volume (in cm<sup>3</sup>) | 2.59(0.69,5.20) | 2.52(0.77,4.91) |
| Six-category scale |  |  |
| 2-Hospitalized, not requiring supplemental oxygen | 14 (21.54%) | 10 (28.57%) |
| 3-Hospitalized, requiring supplemental oxygen | 50 (76.92%) | 25 (71.43%) |
| 4-Hospitalized, on noninvasive ventilation or high flow oxygen devices | 1 (1.54%) | 0 (0.00%) |
| White blood cell count* (10<sup>9</sup>/L) | 5.70(5.00,6.60) | 5.80(5.00,6.80) |
| Lymphocyte count (10<sup>9</sup>/L) | 1.39(1.19,1.80) | 1.47(1.24,1.84) |
| CD4 T cells (/μl) † | 641.00(482.00,760.00) | 734.00(502.00,1031.00) |
| CD8 T cells (/μl) † | 371.00(275.00,520.00) | 401.00(307.00,593.00) |
| B cells (/μl) † | 148.50(99.60,251.00) | 148.50(94.70,248.00) |
| NK cells (/μl) † | 233.50(151.00,393.00) | 197.50(136.00,309.00) |
| Neutrophil count (10<sup>9</sup>/L) | 3.48(2.91,4.32) | 3.83(2.85,4.48) |
| Platelet count (10<sup>9</sup>/L) | 214.00(174.00,255.00) | 210.00(176.00,247.00) |
| Hemoglobin ( g/L ) | 122.68 (14.44) | 124.26 (11.83) |
| D-dimer (mg/L) ‡ | 0.58 (0.36,1.11) | 0.56 (0.31,1.12) |
| IL-6 (pg/ml) § | 7.86(5.63,9.84) | 8.76(6.54,11.77) |
| CRP(mg/L) ¶ | 1.95(0.84,3.53) | 1.38(0.68,2.26) |
| SARS-CoV-2 test result\|\| |  |  |
| SARS-Cov-2 IgG positive | 63 (100.00%) | 34 (100.00%) |
| SARS-Cov-2 IgM positive | 58 (92.06%) | 32 (94.12%) |
| SARS-Cov-2 nucleic acid detection positive | 47(72.31%) | 20(57.14%) |

Data are median (interquartile range (IQR)), n (%),or mean (SD)
\* BMI values were available for 59 patients in the UC-MSC group and 33 patients in the placebo group.
† CD4, CD8, CD19, and CD56 values were available for 62 patients in the UC-MSC group and 34 patients in the placebo group.
‡ D-dimer values were available for 55 patients in the UC-MSC group and 29 patients in the placebo group.
§ IL-6 values were available for 64 patients in the UC-MSC group and 35 patients in the placebo group.
¶ CRP values were available for 27 patients in the UC-MSC group and 14 patients in the placebo group.
|| The test results are summarized from hospitalization to the pre-random test. If there is any positive, it is defined as positive.
The IgG and IgM values were available for 63 patients in the UC-MSCs group and 34 patients in the placebo group.

### Imaging and clinical outcomes

To evaluate the difference in the primary endpoint and parts of the secondary endpoints, we analysed the changes in high-resolution chest CT images and measured the lesions by using centralised imaging interpretation based on the evaluation of both radiologist analyses and lung imaging artificial intelligence software. Through comparison of the Hodges-Lehmann estimator of the total lesion proportion (%) of the whole lung volume, the median change was -19.40% (95% CI, -53.40%, -2.62%) in the MSC group, and -7.30% (95%CI, -46.59%, 19.12%) in the placebo group on day 28 from baseline, yielding a difference of -13.31% (95% CI, -29.14%, 2.13%, *P*= .080) (Figure 2, Table 2, Supplement 5). Interestingly, in the evaluation of the solid component lesions as a specific lesion type, we found that the median change from baseline to day 28 were -57.70% (95%CI, -74.95%, -36.56%) and -44.45% (95% CI, -62.24%, -8.82%) in the MSC and placebo groups, respectively, leading to a significant difference between the MSC group and placebo group (95% CI -30.82%, -0.39%, *P*= .043). We also observed decrease in the ground-glass lesions in the MSC group than the placebo group, although the difference was not statistically significant. To exclude the effect of missing data on the outcomes of the above results in some mITT cases, we further analysed the PP population. Almost identical results between the two groups were obtained, as shown in Supplement 6 and 7.

**Table 2:**
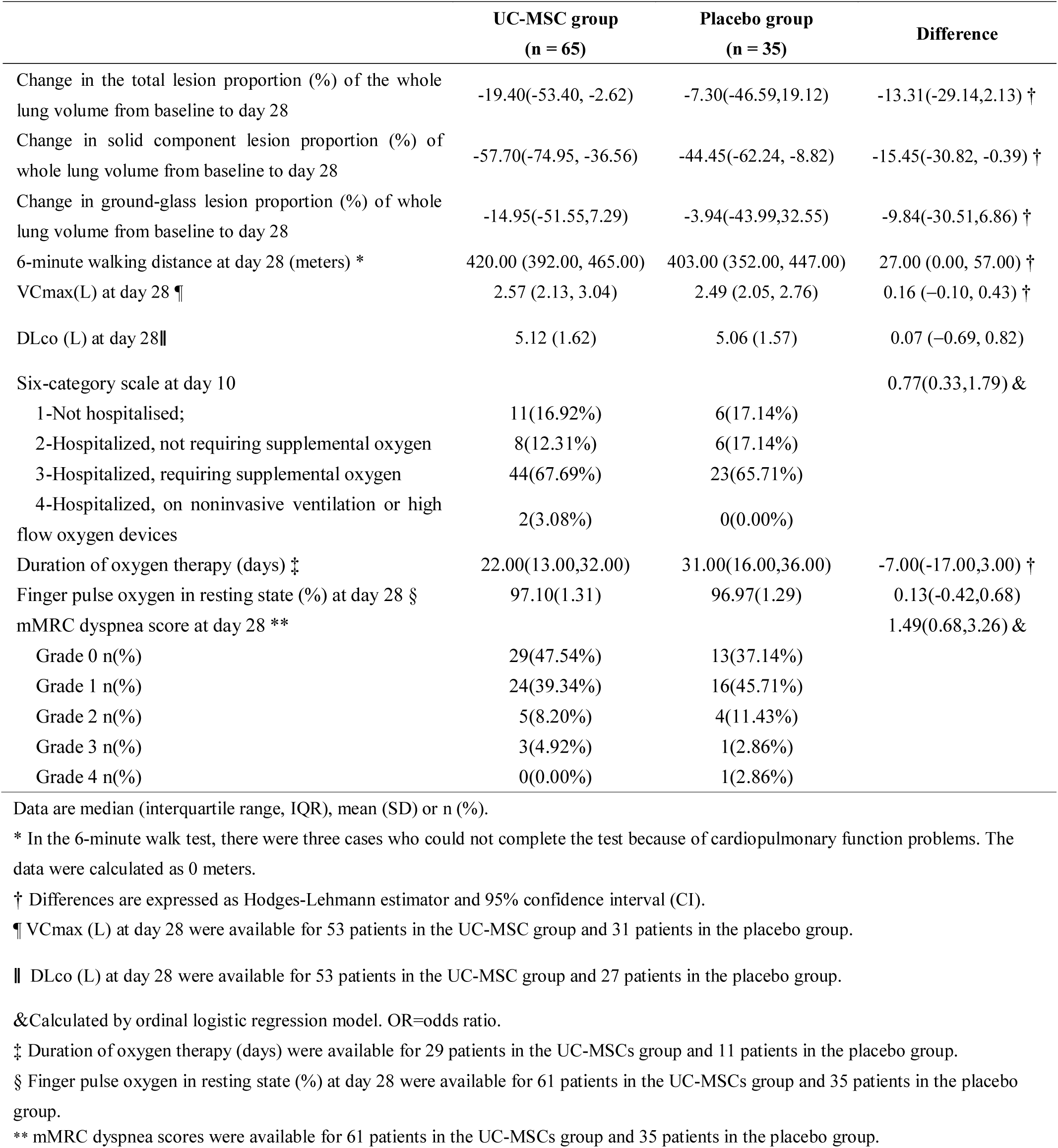
Primary and secondary outcomes in the mITT population.

|  | UC-MSc group<br>(n = 65) | Placebo group<br>(n = 35) | Difference |
| --- | --- | --- | --- |
| Change in the total lesion proportion (%) of the whole lung volume from baseline to day 28 | -19.40(-53.40, -2.62) | -7.30(-46.59,19.12) | -13.31(-29.14,2.13) † |
| Change in solid component lesion proportion (%) of whole lung volume from baseline to day 28 | -57.70(-74.95, -36.56) | -44.45(-62.24, -8.82) | -15.45(-30.82, -0.39) † |
| Change in ground-glass lesion proportion (%) of whole lung volume from baseline to day 28 | -14.95(-51.55,7.29) | -3.94(-43.99,32.55) | -9.84(-30.51,6.86) † |
| 6-minute walking distance at day 28 (meters) * | 420.00 (392.00, 465.00) | 403.00 (352.00, 447.00) | 27.00 (0.00, 57.00) † |
| VCmax(L) at day 28 ¶ | 2.57 (2.13, 3.04) | 2.49 (2.05, 2.76) | 0.16 (-0.10, 0.43) † |
| DLco (L) at day 28 \|\| | 5.12 (1.62) | 5.06 (1.57) | 0.07 (-0.69, 0.82) |
| Six-category scale at day 10 |  |  | 0.77(0.33,1.79) & |
| 1-Not hospitalised; | 11(16.92%) | 6(17.14%) |  |
| 2-Hospitalized, not requiring supplemental oxygen | 8(12.31%) | 6(17.14%) |  |
| 3-Hospitalized, requiring supplemental oxygen | 44(67.69%) | 23(65.71%) |  |
| 4-Hospitalized, on noninvasive ventilation or high flow oxygen devices | 2(3.08%) | 0(0.00%) |  |
| Duration of oxygen therapy (days) ‡ | 22.00(13.00,32.00) | 31.00(16.00,36.00) | -7.00(-17.00,3.00) † |
| Finger pulse oxygen in resting state (%) at day 28 § | 97.10(1.31) | 96.97(1.29) | 0.13(-0.42,0.68) |
| mMRC dyspnea score at day 28 ** |  |  | 1.49(0.68,3.26) & |
| Grade 0 n(%) | 29(47.54%) | 13(37.14%) |  |
| Grade 1 n(%) | 24(39.34%) | 16(45.71%) |  |
| Grade 2 n(%) | 5(8.20%) | 4(11.43%) |  |
| Grade 3 n(%) | 3(4.92%) | 1(2.86%) |  |
| Grade 4 n(%) | 0(0.00%) | 1(2.86%) |  |

Data are median (interquartile range, IQR), mean (SD) or n (%).
\* In the 6-minute walk test, there were three cases who could not complete the test because of cardiopulmonary function problems. The data were calculated as 0 meters.
† Differences are expressed as Hodges-Lehmann estimator and 95% confidence interval (CI).
¶ VCmax (L) at day 28 were available for 53 patients in the UC-MSc group and 31 patients in the placebo group.
|| DLco (L) at day 28 were available for 53 patients in the UC-MSc group and 27 patients in the placebo group.
&Calculated by ordinal logistic regression model. OR=odds ratio.
‡ Duration of oxygen therapy (days) were available for 29 patients in the UC-MSCs group and 11 patients in the placebo group.
§ Finger pulse oxygen in resting state (%) at day 28 were available for 61 patients in the UC-MSCs group and 35 patients in the placebo group.
\*\* mMRC dyspnea scores were available for 61 patients in the UC-MSCs group and 35 patients in the placebo group.

**Figure 2.**
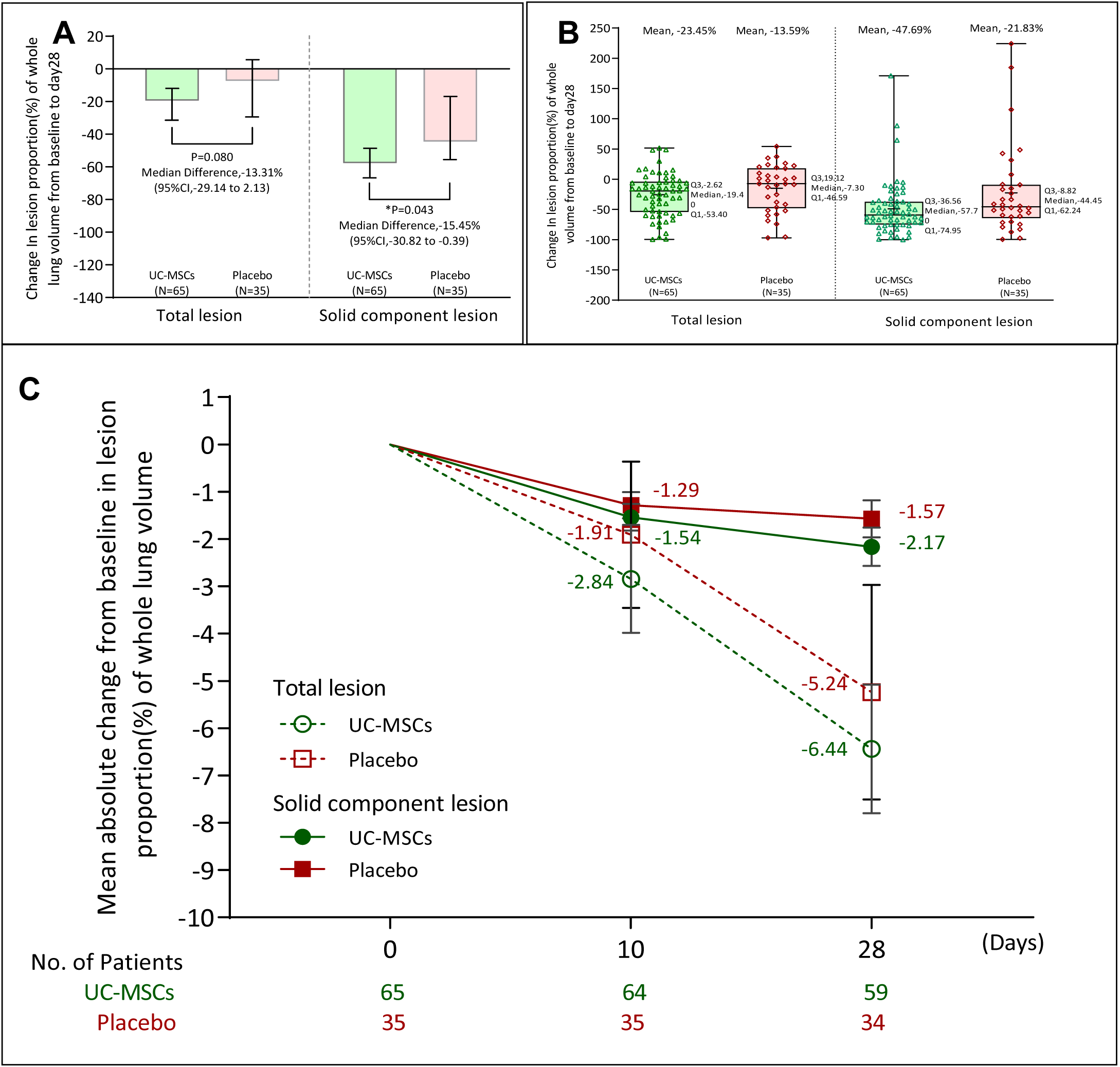
Decline in total and solid component lesion proportion (%) between the MSC group and placebo group at 28 day. A. Panel A shows the between-group median difference in the change in total lesion proportion (%) and solid component lesion proportion (%) of the whole lung volume from baseline to day 28. I bars indicate the 95%CI described by Hahn and Meeker (1991). B. Panel B shows box plots of the change in the total lesion proportion (%) and solid component lesion proportion (%) of the whole lung volume from baseline to day 28. Q1 denotes the first quartile, and Q3 the third quartile. I bars indicate the minimum and maximum. C. Panel C shows the mean absolute change from baseline in the total lesion proportion (%) and solid component lesion proportion (%) of the whole lung volume. I bars indicate the standard error.

To compare the restoration of lung function and integrated reserve capability among the two groups of patients, we examined the 6-MWT on the 28th day after the onset of treatment and found that 6-minute walking distance was longer in the MSC group (median 420.00 meters [interquartile range (IQR) 392.00,465.00]) than in the placebo group (median 403.00 meters [IQR 352.00,447.00]) with a 95% CI of 0.00–57.00 (*P*= .057, Table 2). Other parameters including VCmax and DLco, the six-category scale, status of oxygen therapy and mMRC dyspnoea score were similar between the two groups (Table 2). In addition, there was no significant difference in the subsets of peripheral lymphocyte counts (CD4+ T cells, CD8+ T cells, B cells, NK cells) and plasma markers between the two groups (Supplement 8).

### Post Hoc Analyses

We have established five models in three analysis data sets (mITT, PPS and ITT) for sensitivity analyses of primary end point (Supplement 9). The five models included: treatment group factor was included in model 1, treatment group and center factors were included in model 2 as fixed effects, and treatment group, baseline and center factors were included as fixed effects in model 3, center factor was included as random effect, treatment group factor was included as fixed effects in model 4, center factor was included as random effect, treatment group and baseline factors were included as fixed effects in model 5. The conclusions of all models were consistent with those of univariate analysis.

### Safety

The incidence of adverse events reported during the study was similar in the MSC group (55.38%) and the placebo group (60%) (Table 3). The most common adverse event in the MSC group was an increase in lactic acid dehydrogenase (13.85%), compared with 20% in the placebo group; a 10.77% elevation of serum alanine aminotransferase compared with 11.43% in the placebo group; a 9.23% increase in hypokalaemia compared with 2.86% in the placebo group; a 7.69% increase in aspartate aminotransferase compared with 11.43% in the placebo group; and a 7.69% increase in hyperuricemia compared with 8.75% in the placebo group. Only one case experienced a grade 3 adverse event (pneumothorax) in the MSC group, which recovered spontaneously under conservative treatment. There were few other adverse events at grade 1 or 2 in both groups. All adverse events during the observation period were judged by the site investigators and found to be unrelated to UC-MSC intervention. No deaths were observed in this trial.

**Table 3.** Summary of adverse events that occurred in the enrolled population of the trial.

|  | UC-MSD group (n = 65) |  | Placebo group (n = 35) |  |
| --- | --- | --- | --- | --- |
|  | Grade 1 or 2 | Grade 3 or 4 | Grade 1 or 2 | Grade 3 or 4 |
| **Any adverse event** | 36 (55.38%) | 1 (1.54%) | 21 (60.00%) | 0 (0.00%) |
| Increased lactic acid dehydrogenase | 9(13.85%) | 0(0.00%) | 7(20.00%) | 0(0.00%) |
| Increased alanine aminotransferase | 7(10.77%) | 0(0.00%) | 4(11.43%) | 0(0.00%) |
| Hypokalaemia | 6(9.23%) | 0(0.00%) | 1(2.86%) | 0(0.00%) |
| Increased aspartate aminotransferase | 5(7.69%) | 0(0.00%) | 4(11.43%) | 0(0.00%) |
| Increased serum uric acid | 5(7.69%) | 0(0.00%) | 3(8.57%) | 0(0.00%) |
| Diarrhoea | 4(6.15%) | 0(0.00%) | 0(0.00%) | 0(0.00%) |
| Palpitations | 3(4.62%) | 0(0.00%) | 0(0.00%) | 0(0.00%) |
| Increased $\gamma$ -glutamyl transferase | 2(3.08%) | 0(0.00%) | 1(2.86%) | 0(0.00%) |
| Dizziness | 2(3.08%) | 0(0.00%) | 0(0.00%) | 0(0.00%) |
| Cough | 2(3.08%) | 0(0.00%) | 1(2.86%) | 0(0.00%) |
| abdominal distention | 2(3.08%) | 0(0.00%) | 0(0.00%) | 0(0.00%) |
| Anemia | 2(3.08%) | 0(0.00%) | 0(0.00%) | 0(0.00%) |
| Pneumothorax | 0(0.00%) | 1(1.54%) | 0(0.00%) | 0(0.00%) |
| Metabolic alkalosis | 1(1.54%) | 0(0.00%) | 0(0.00%) | 0(0.00%) |
| Urinary tract infection | 1(1.54%) | 0(0.00%) | 0(0.00%) | 0(0.00%) |
| Bacterial infection | 1(1.54%) | 0(0.00%) | 0(0.00%) | 0(0.00%) |
| Pharyngitis | 1(1.54%) | 0(0.00%) | 0(0.00%) | 0(0.00%) |
| Increased heart rate | 1(1.54%) | 0(0.00%) | 0(0.00%) | 0(0.00%) |
| Creatine phosphate stimulation | 1(1.54%) | 0(0.00%) | 1(2.86%) | 0(0.00%) |
| Elevated blood urea | 1(1.54%) | 0(0.00%) | 1(2.86%) | 0(0.00%) |
| Poor sleep quality | 1(1.54%) | 0(0.00%) | 0(0.00%) | 0(0.00%) |
| Taste reversal | 1(1.54%) | 0(0.00%) | 0(0.00%) | 0(0.00%) |
| Chest musculoskeletal | 1(1.54%) | 0(0.00%) | 0(0.00%) | 0(0.00%) |
| Pulmonary edema | 1(1.54%) | 0(0.00%) | 0(0.00%) | 0(0.00%) |
| Pharyngeal diseases | 1(1.54%) | 0(0.00%) | 0(0.00%) | 0(0.00%) |
| Anxious | 1(1.54%) | 0(0.00%) | 0(0.00%) | 0(0.00%) |
| Nervous | 1(1.54%) | 0(0.00%) | 0(0.00%) | 0(0.00%) |
| Rash | 1(1.54%) | 0(0.00%) | 0(0.00%) | 0(0.00%) |
| Thirsty | 1(1.54%) | 0(0.00%) | 0(0.00%) | 0(0.00%) |
| Nausea | 1(1.54%) | 0(0.00%) | 0(0.00%) | 0(0.00%) |
| Countercurrent | 1(1.54%) | 0(0.00%) | 0(0.00%) | 0(0.00%) |
| Non-infectious gingivitis | 1(1.54%) | 0(0.00%) | 0(0.00%) | 0(0.00%) |
| Abdominal pain | 1(1.54%) | 0(0.00%) | 0(0.00%) | 0(0.00%) |
| Functional gastrointestinal turbulence | 1(1.54%) | 0(0.00%) | 0(0.00%) | 0(0.00%) |
| Vomit | 1(1.54%) | 0(0.00%) | 0(0.00%) | 0(0.00%) |
| Gastroesophageal reflux disease | 1(1.54%) | 0(0.00%) | 1(2.86%) | 0(0.00%) |
| Toothache | 1(1.54%) | 0(0.00%) | 0(0.00%) | 0(0.00%) |
| heart failure | 1(1.54%) | 0(0.00%) | 0(0.00%) | 0(0.00%) |
| Hypocalcemia | 0(0.00%) | 0(0.00%) | 2(5.71%) | 0(0.00%) |
| Hepatic cyst | 0(0.00%) | 0(0.00%) | 2(5.71%) | 0(0.00%) |
| Creatine phosphate stimulation | 0(0.00%) | 0(0.00%) | 1(2.86%) | 0(0.00%) |
| Elevated serum creatinine | 0(0.00%) | 0(0.00%) | 1(2.86%) | 0(0.00%) |
| Respiratory alkalosis | 0(0.00%) | 0(0.00%) | 1(2.86%) | 0(0.00%) |
| Pleural effusion | 0(0.00%) | 0(0.00%) | 1(2.86%) | 0(0.00%) |
| Difficulty in falling asleep | 0(0.00%) | 0(0.00%) | 1(2.86%) | 0(0.00%) |
| Pruritus | 0(0.00%) | 0(0.00%) | 3(8.57%) | 0(0.00%) |

## Discussion

Whilst several trials of the therapeutic use of MSCs for patients with COVID-19 have been registered at Clinicaltrial.gov, there are no data available to date from randomized placebo-controlled clinical trials yet. This is the first report of a double-blind, randomized, and controlled phase 2 trial, in COVID-19 patients with lung damage. Our data found that UC-MSC administration was safe and well tolerated and exerted a trend of improvement in whole lung lesion for COVID-19 patients. More interestingly, UC-MSC medication significantly increased the resolution of lung solid component lesions compared with the placebo. The data from the 6-MWT data show an improved restoration of the integrated reserve capability in the UC-MSC-treated patients. These findings indicate that the use of UC-MSC as adjunctive therapy to standard of care treatment for patients with COVID-19 is a viable option. Recently, MSCs have been approved conditionally by the US-FDA under what is known as ‘expanded access compassionate use’ for COVID-19 patients. A phase 3 trial is now required to further evaluate effects on mortality and long term pulmonary disability, and to determine the underlying mechanisms of UC-MSC treatment for COVID-19 disease.^22^

COVID-19 is characterized by pathological lung changes in both the parenchyma and interstitium^2^, including ground glass opacity, solid component, traction bronchiectasis, reticulation, and thickening of the bronchovascular bundles, as imaged using chest CT^23^. Notably, the improvement of pulmonary lesions, especially the improvement of pulmonary interstitial lesions, directly affects the recovery of lung function and the remission of clinical symptoms.^24^ The outbreak of COVID-19 occurred in China in January and early February, and was basically brought under control in March. As enrolment ensured, many severe hospitalized COVID-19 patients were at the convalescent stage and some of these patients in the progression stage, which presented a challenge to explore clinical improvement. Considering lung damage was still a common characteristic in these patients at convalescent phase which affected their recovery and life quality, we modified the primary outcome to the change in the total lesion proportion (%) of the whole lung volume as measured by CT from baseline to day 28. The patients enrolled in this study were all previously diagnosed as severe types, with a longer disease course and older age, compared with the recent studies.^25,26^ In particular, all the patients suffered from serious pulmonary damage and needed oxygen inhalation support during the course of disease. Our current trial showed that UC-MSCs therapy improved the resolution of the whole lung damage size, as detected by CT scanning, particularly the solid component lesions. This finding indicated that UC-MSC administration has a therapeutic benefit for patients with COVID-19, even in the convalescent stage. It is known that the solid component lesions in the lung include the interstitial fibrosis. Thus, the improvements of solid component lesions might also imply the alleviation of lung fibrosis.

The 6-MWT has been used to evaluate patients suffering from a variety of cardiopulmonary diseases. The results reflect the integrated reserve capability of complex physiology, involving the pulmonary and cardiovascular systems, and neuromuscular circulation^27^. In this trial, the 6-MWT was numerically, but not statistically, improved in the MSC group compared with that in the placebo group. Given that there was no significant difference in cardiovascular diseases between the two groups and no cerebrovascular diseases in both groups at baseline, the findings from the 6-MWT data imply that capacity for aerobic exercise was potentially improved in the UC-MSCs group.

It is hypothesized that the beneficial effect of MSC treatment for patients with severe COVID-19 is mediated via reduction of pro-inflammatory cytokines, that jointly mediate immune pathology and worsen clinical COVID-19 outcomes.^7,28-30^ Cytokines such as serum IL-6 are considered as biologically relevant biomarkers associated with disease progression of COVID-19. In this study, UC-MSC infusion did not result in a significant reduction in the duration of oxygen therapy, mMRC, cytokine, or chemokine levels, which might be in part attributed to the status of the enrolled population, since most of them were not in the acute progressive stage. Other mechanisms of actions have to be explored for MSCs that are measurable in the systemic circulation. We could not, however, measure the local, i.e. intrapulmonary, effects of MSC delivery. It could very well be that the local MSC-mediated effects were not measurable in the systemic circulation, a similar scenario as in MSC treatment of patients with corticosteroid-resistant graft-versus-host-disease (GVHD).

In our trial, a total of three doses of 4 × 10^7^ UC-MSCs were transfused for each patient. No MSC-related predefined haemodynamic or respiratory adverse events were observed. The incidences of adverse events were similar between the MSC group and the placebo group. Only one patient in the MSC group suffered a pneumothorax that was judged to be unrelated to UC-MSC medication. No patient died during the follow-up period. The safety profile of the UC-MSCs confirms the results of our previous phase 1 trial^20^ and other MSC studies.^31,32^ These data suggested that UC-MSC therapy was well tolerated and very safe. In view of the ongoing clinical trials with higher dose of MSCs used in other groups in China, America, and Europe,^22^ more safety data are expected in the near future.

There were several operational limitations to our study. A larger sample size could have improved efficacy analyses. According to management guidelines issued by the Chinese National Health Commission (7^th^ edition),^21^ patients with COVID-19 require further centralized isolation for 14 days after discharge. In this setting, some of the patients missed the follow-up data at day 28, but they did receive a follow-up check around 7–10 days after the 28-day follow-up window. Importantly, our PP population analysis also revealed similar results compared with mITT population analysis. Whether the cell dosage, interval duration, and cycles of UC-MSC medication were the best regimen for patients with severe COVID-19 were not fully investigated in this study.

To the best of our knowledge, this is the first randomized, double-blind, placebo-controlled trial evaluating the safety and preliminary efficacy of UC-MSCs as a potential treatment for patients with COVID-19 with lung damage, even at the convalescent stage. UC-MSC administration was safe and accelerated resolution of lung solid component lesions and improvement in the integrated reserve capability after UC-MSC administration. UC-MSCs treatment offers a safe and potentially effective therapeutic approach for COVID-19 patients with lung damage. A phase 3 trial is required to further evaluate effects on preventing long-term pulmonary disability, reducing mortality and determining the underlying mechanisms of UC-MSC treatment for COVID-19 disease.

## Supporting information

Supplement 1-Protocol-V 3.1 Final

Supplement 2-SAP

## Data Availability

All data referred in the manuscript are available on resonable request to Fu-Sheng Wang at.
Additional information
Who can access the data: researchers whose proposed use of the data has been approved
Types of analysis: Such requests are at the Academic Committee's discretion and dependent on the nature of the request, the merit of the research proposed, availability of the data and the intended use of the data.
Mechanisms of data availability: After approval from the Academic Committee, this trial data can be shared with qualifying researchers who submit a proposal with a valuable research question. A contract should be signed.
Any additional restrictions: NA.

## Contributions

FSW ideated and led the study. FSW, WFX, CY and LS designed the study and developed the protocol. LS, HH, XCL, XY, ZX, LH, JLF and LLS were responsible for study enrolment. LS, XCL, XY, ZX, LH, JLF were responsible for acquisition, analysis, and interpretation of data. YZ, WQY, CZ, TYL and JWS were responsible for biorepository management and biomarker analyses. All authors made substantial contributions to conduct and coordination of trial and had regular discussions on progress of the study. CY, YXY and YPY contributed to statistical analysis. LS, RNX and CZ wrote the initial manuscript draft, and FSW, WFX, MM and AZ critically revised the manuscript. All authors edited, read and approved the final version before submission.

## Ethical approval

This study was approved by the Clinical Trial Ethics Committee of Fifth Medical Cente, Chinese PLA General Hospital (2020-013-D), the Medical Ethics Committee of Wuhan Huoshenshan Hospital(HSSLL004), the Medical Ethics Committee of Maternal and Child Hospital of Hubei Province(2020IEC001).Written informed consent was obtained from all the enrolled patients or their legal representatives if they were unable to provide consent.

## Data sharing

After approval from the Human Genetic Resources Administration of China, this trial data can be shared with qualifying researchers who submit a proposal with a valuable research question. A contract should be signed.

## Acknowledgements

F-S.W, C.Z, M.M., and A.Z. are founder members of the Global Cancer and Infectious Diseases consortium for Host-Directed therapies (https://www.fchampalimaud.org/covid19/aci/)

## Additional Contributions

We thank Drs. Sibing Zhang, Dixiong Xu, Fangguo Dai and Bin Liu for logistic support and valuable suggestions to the clinical study. We also thank Fang Lian, Baoqiang Fu, Yingying Gao and Yan Zhang for their excellent work in follow-up of all enrolled patients. We thank randomization system provider (Chengdu Cims-medtech), PIATS provider (Alibaba Health Technology) and academic secretaries (Yufeng Guo, Hairui Si, Lvshuai Huang, Junqing Luan) for their services. We also thank the People’s Government of Wuhan Municipality for the help in organizing the follow-up work.

