## Supplement 1-Protocol-V 3.1 Final for "Treatment with human umbilical cord-derived mesenchymal stem cells for COVID-19 patients with lung damage: a randomised, double-blind, placebo-controlled phase 2 trial"

### A phase II, multicenter, randomized, double-blind, placebo-controlled trial to evaluate the efficacy and safety of human umbilical cord-derived mesenchymal stem cells in the treatment of severe COVID-19 patients

#### Protocol

Version 3.1 - June 1, 2020

| | |
|---|---|
| Sponsor: | The Fifth Medical Center of PLA General Hospital |
| Principal investigator: | Fu-Sheng Wang |
| Ethics Reference Number: | 2020-013-D |
| ClinicalTrials.gov | NCT04288102 |

#### 1. TRIAL SUMMARY

World Health Organization Registration Data Set

| | |
|---|---|
| Title | A phase II, multicenter, randomized, double-blind, placebo-controlled trial to evaluate the efficacy and safety of human umbilical cord-derived mesenchymal stem cells in the treatment of severe COVID-19 patients |
| Primary registry and trial identifying number | ClinicalTrials.gov NCT04288102 |
| Secondary identifying numbers | 2020-013-D |
| Sources of monetary or material support | The Fifth Medical Center of PLA General Hospital |
| Primary sponsor | The Fifth Medical Center of PLA General Hospital |
| Central contact | Principal Investigator: Fu-Sheng Wang, MD, Ph.D.<br>86-10-66933332<br><br>Investigator and coordinator: Lei Shi, MD, Ph.D.<br>86-10-66933333<br><br>Study Sites:<br><br>Wuhan Huoshenshan Hospital, Wuhan, Hubei, China, 430000<br>Contact: Lei Shi, MD, PhD<br><br>Optical Valley Branch of Maternal and Child Hospital of Hubei Province, Wuhan, Hubei, China, 430000<br>Contact: Wei-Fen Xie, MD, PhD<br><br>General Hospital of Central Theater Command (Follow-up only), Hubei, China, 430000<br>Contact: Xiaojing Jiang, MD, PhD |
| Study officials/Investigators | Study Principal Investigator<br>Fu-Sheng Wang, MD, PhD<br>The Fifth Medical Center of PLA General Hospital |
| Brief title | Treatment with human umbilical cord-derived mesenchymal stem cells for severe corona virus disease 2019 (COVID-19) |

| | |
|---|---|
| Countries of recruitment | P. R. China |
| Condition(s) or focus of study | COVID-19 |
| Interventions | Human umbilical cord-derived mesenchymal stem cells (UC-MSCs) |
| Key eligibility criteria | **Inclusion criteria**<br>1. Age≥18 and <75 years;<br>2. hospitalized;<br>3. Severe COVID-19:<br>1) Laboratory confirmation of SARS-CoV-2 infection by reverse-transcription polymerase chain reaction (RT-PCR) from any diagnostic sampling source<br>2) Confirmed pneumonia by chest computed tomography imaging<br>3) Apply to any one of the following: 1)dyspnea (RR ≥ 30 times/min), 2)finger oxygen saturation ≤ 93% in resting state, 3)arterial oxygen partial pressure (PaO2) / oxygen absorption concentration (FiO2) ≤ 300MMHG, 4)pulmonary imaging shows that the focus progress > 50% in 24-48 hours<br>4. Confirmed interstitial lung damage by chest computed tomography imaging.<br>**Exclusion criteria**<br>1. Pregnancy, lactation and those who are not pregnant but do not take effective contraceptives measures;<br>2. Patients with malignant tumor, other serious systemic diseases and psychosis;<br>3. Patients who are participating in other clinical trials;<br>4. Inability to provide informed consent or to comply with test requirements.<br>5. Co-Infection of HIV, tuberculosis, influenza virus, adenovirus and other respiratory infection virus.<br>6. Invasive ventilation<br>7. Shock<br>8. Combined with other organ failure( need organ support)<br>9. Interstitial lung damage caused by other reasons ( in 2 weeks) |

| | |
|---|---|
| | 10. The pulmonary imaging revealed the interstitial damage of lungs before the COVID-19 confirmed. |
| Study design | Study type: Interventional trial<br>Allocation: Randomized<br>Intervention model: Parallel group<br>Primary purpose: Treatment<br>Phase: Phase 2 |
| Masking | Double-blind |
| Date of enrollment | March 5, 2020. |
| Target sample size | 90 (60 in UC-MSCs group, 30 in placebo group) |
| Recruitment status | Recruiting |
| Primary outcomes | **Primary endpoint measures (**centralized imaging interpretation**)**<br>• Change in lesion proportion (%) of full lung volume from baseline to day 28.<br>Lesion proportion = lesion volume (in $cm^3$) / full lung volume (in $cm^3$) |
| Secondary outcomes | **Secondary endpoint measures (**centralized imaging interpretation**)**<br>• Change in lesion proportion (%) of full lung volume from baseline to day 10 and 90<br>• Change in solid component lesion proportion (%) of full lung volume from baseline to day 10, 28 and 90.<br>• Change in ground-glass lesion proportion (%) of full lung volume from baseline to day 10, 28 and 90.<br>• Pulmonary fibrosis – related morphological features in CT scan at day 90<br>a. cord-like shadow<br>b. honeycomb-like shadows<br>c. interlobular septal thickening<br>d. intralobular interstitial thickening<br>e. pleural thickening<br>• Lung densitometry:<br>a. Change in total voxel ‘weight’ in lesion area from baseline to day 10, 28 and 90.<br>voxel ‘weight’=voxel density (in HU) × voxel volume (in voxel) |

b. volumes histogram of lung density distribution (<-750, -750~-300, -300~50, >50) at day 10, 28 and 90.

**Other secondary endpoints**

- Time to clinical improvement in 28 days.

  Clinical improvement defined as a one-point deduction from baseline in a 6 ordinal scale:

  1. Not hospitalized;
  2. Hospitalized, not requiring supplemental oxygen;
  3. Hospitalized, requiring supplemental oxygen;
  4. Hospitalized, on non-invasive ventilation or high flow oxygen devices;
  5. Hospitalized, on invasive mechanical ventilation or ECMO;
  6. Death.
- Change in oxygenation index ($PaO2/FiO2$) from baseline to day 6, 10, and 28.
- The duration of oxygen therapy (in days)
- Change in oxygen saturation from baseline to day 6, 10 and 28
- 6-minute walk test at day 28 and 90 (in meters)
- Pulmonary function test at day 28 and 90
  a. maximum vital capacity ($VC_{max}$)
  b. diffusing Capacity ($DL_{CO}$)
- Difference in the change of mMRC (Modified Medical Research Council) Dyspnea Scale at day 28, day 90.
- Changes of absolute lymphocyte counts and subsets, as well as cytokine/chemokine levels from baseline to day 6, 10, 28 and 90.

### 2. INTRODUCTION

#### 2.1. Background and rationale

The Corona Virus Disease 2019 (COVID-19) caused by severe acute respiratory syndrome corona virus 2 (SARS-CoV-2) infection has unprecedentedly spread in the worldwide and been declared as a pandemic by the world health organization. COVID-19 is characterized by sustained cytokines production and hyper-inflammation, can cause clusters of severe respiratory illness with a fatality rate around 2-5%. There are currently no prophylactic vaccine and no specific antiviral treatment agents available recommended for COVID-19. The management of COVID-19 patients remains largely symptomatic and supportive therapy for severe patients. Therefore, it is urgent to find a safe and effective therapeutic approach to COVID-19.

During the last decade, the promising features of mesenchymal stem cells (MSCs), including their regenerative properties and ability to differentiate into diverse cell lineages, have generated great interest among researchers whose work has offered intriguing perspectives on cell-based therapies for various diseases. MSCs have already secured conditional approved for the treatment of children with graft-versus-host disease for its inflammation suppressing previlege. MSCs could significantly reduce the pathological changes of lung and inhibit the cell-mediated immune inflammatory response induced by influenza virus in animal model and clinical trial. In previous study, the safety and primary efficacy of MSC have been evaluated in patients with acute respiratory distress syndrome (ARDS) and the underlying mechanisms have been studied in some pre-clinical settings. It seems that MSC-based treatments principally contributed to inhibit the uncontrolled immune inflammatory response, benefit the recovery of lung function, and were conductive to delay the progress of lung fibrosis. Accordingly, these findings raise the hypothesis that use of MSCs transfusion could be beneficial in COVID-19 patients.

As a first step in testing safety, we did a phase 1, open-label trial in 20 patients with severe COVID-19, using a dose of $3\times10^7$ umbilical cord-MSCs(UC-MSCs) at 3-day intervals by intravenous reinfusion that may contribute to alleviate hyper-inflammation response. Infusions were well tolerated. Given the in-vitro and in-vivo benefit of MSCs and the acceptable tolerance and safety in humans, we developed this trial with the objective of evaluating the safety and efficacy of intravenous UC-MSCs in adults with severe pneumonia caused by SARS-CoV-2.

#### 2.2. Objectives

To evaluate the efficacy and safety of umbilical cord-derived human mesenchymal stem cells in the treatment of severe COVID-19 patients.

#### 2.3. Trial design

This trial is designed as a phase II, multi-center, randomized, double-blind, placebo-controlled trial. The allocation ratio is 2:1 in the testing group comparing to the control group. This is an investigator-initiated trial.

### 3. METHODS

#### 3.1. Study setting

The trial will be conducted in two COVID-19 designated hospitals in Wuhan City, Hubei Province: Huoshenshan Hospital and Hubei Maternity and Child Health Care Hospital. Huoshenshan Hospital is a new-built hospital for this outbreak with 1000 beds capacity, and Hubei Maternity and Child Health Care Hospital is temporarily reformed for admitting COVID-19 patients from an under constructed specialized hospital with 800 beds capacity. After patients discharged from the COVID-19 designated hospitals, the follow-up procedure will be conducted at the General Hospital of Central Theater Command of PLA.

#### 3.2. Eligibility criteria

##### *3.2.1. Inclusion criteria*

1. Age ≥18 and <75 years;
2. hospitalized;
3. Severe COVID-19:

    1) Laboratory confirmation of SARS-CoV-2 infection by reverse-transcription polymerase chain reaction (RT-PCR) from any diagnostic sampling source
    2) Confirmed pneumonia by chest computed tomography imaging
    3) Apply to any one of the following after onset: 1)dyspnea (RR ≥ 30 times/min), 2)finger oxygen saturation ≤ 93% in resting state, 3)arterial oxygen partial pressure (PaO2) / oxygen absorption concentration (FiO2) ≤ 300MMHG, 4)pulmonary imaging shows that the focus progress > 50% in 24-48 hours

4. Confirmed interstitial lung damage by chest computed tomography imaging.

##### *3.2.2. Exclusion criteria*

1. Pregnancy, lactation and those who are not pregnant but do not take effective contraceptives measures
2. Patients with malignant tumor, other serious systemic diseases and psychosis
3. Patients who are participating in other clinical trials
4. Inability to provide informed consent or to comply with test requirements.
5. Co-Infection of HIV, tuberculosis, influenza virus, adenovirus and other respiratory infection virus.
6. Invasive ventilation
7. Shock
8. Combined with other organ failure( need organ support)
9. Interstitial lung damage caused by other reasons ( in 2 weeks)

10. The pulmonary imaging revealed the interstitial damage of lungs before the COVID-19 confirmed.

#### 3.3. Interventions

##### *3.3.1. Investigational medicinal product*

In this study, the umbilical cord-derived mesenchymal stem cells (UC-MSCs) will be tested for treatment. The UC-MSCs are harvested from human umbilical cord. The product is almost colorless suspension, which contains $4.0 \times 10^7$ MSCs with a volume of 100ml/bag. The placebos are with the same appearance in packaging and the suspension without MSCs. The dose of the treatment is $4.0 \times 10^7$ (1 bag) / procedure, three procedures for every patient on day 0, day 3, and day 6 after randomization.

The administration procedure is as follows:

1) check the patient status and emergency equipment
2) verify UC-MSC packaging and patient information
3) use saline to flush the catheter before infusion
4) shake the bag gently when flocculant is notable in the bag
5) intravenous infusions in 30 minutes, press the bag gently to prevent cell clumping if necessary.
6) use 10 ml to 30 ml saline to flush the bag to ensure maximum MSC infusion
7) closely monitoring the patients' ECG, blood oxygen saturation, body temperature, pulse, skin color, respiration, and blood pressure during the entire procedure.
8) Dispose of medical waste with protection to prevent potential virus transmission

##### *3.3.2. Standard of care (SOC)*

The standard of care is following the updated guideline issued by the Chinese National Health Commission (7$^{th}$ edition). The principle of the treatment is supportive care in addition to infection prevention and control as necessary, including the following measures: respirational support (supplemental oxygen, noninvasive and invasive ventilation, and extracorporeal membrane oxygenation), vasopressor support, renal-replacement therapy.

Any concomitant care (medications, procedures) in this study will be recorded.

##### *3.3.3. Intervention assignment*

Experimental group: UC-MSCs administration on day 0, day 3, day 6, plus standard of care during the entire trial.
Control group: UC-MSCs placebo administration on day 0, day 3, day 6, plus standard of care during the entire trial.

Patients will be randomly assigned to either one of the treatment groups. See section 3.8 in detail.

##### *3.3.4. Modifications of intervention*

Criteria for discontinuing of the intervention:

1) Patient withdraws consent
2) Poor compliance results in incomplete administration of UC-MSCs per protocol.
3) Serious adverse events judged by the investigator that the patient is no longer safe to continue UC-MSCs treatment.
4) Any other event judged by the investigator that the patient should be withdrawn from the study

#### 3.4. Outcomes

In this study, the primary endpoint and a part of secondary endpoints will be evaluated by the changes in high-resolution chest computed tomography and measured by a centralized imaging interpretation process.

##### *3.4.1 Imaging Endpoints by centralized interpretation*

**Primary endpoint measures (**centralized imaging interpretation**)**

- Change in lesion proportion (%) of full lung volume from baseline to day 28.
  Lesion proportion = lesion volume (in $cm^3$) / full lung volume (in $cm^3$)

**Secondary endpoint measures (**centralized imaging interpretation**)**

- Change in lesion proportion (%) of full lung volume from baseline to day 10 and 90
- Change in solid component lesion proportion (%) of full lung volume from baseline to day 10, 28 and 90.
- Change in ground-glass lesion proportion (%) of full lung volume from baseline to day 10, 28 and 90.
- Pulmonary fibrosis – related morphological features in CT scan at day 90
  a. cord-like shadow
  b. honeycomb-like shadows
  c. interlobular septal thickening
  d. intralobular interstitial thickening
  e. pleural thickening
- Lung densitometry:
  a. Change in total voxel 'weight' in lesion area from baseline to day 10, 28 and 90.
  voxel 'weight'=voxel density (in HU) × voxel volume (in voxel)
  b. volumes histogram of lung density distribution (<-750, -750~-300, -300~50, >50) at day 10, 28 and 90.

##### *3.4.2 Other secondary endpoints*

1) Time to clinical improvement in 28 days.
   Clinical improvement defined as a one-point deduction from baseline in a 6 ordinal scale:

1. Not hospitalized;
2. Hospitalized, not requiring supplemental oxygen;
3. Hospitalized, requiring supplemental oxygen;
4. Hospitalized, on non-invasive ventilation or high flow oxygen devices;
5. Hospitalized, on invasive mechanical ventilation or ECMO;
6. Death.

2) Change in oxygenation index (PaO2/FiO2) from baseline to day 6, 10, and 28.
3) The duration of oxygen therapy (in days)
4) Change in oxygen saturation from baseline to day 6, 10 and 28
5) 6-minute walk test at day 28 and 90 (in meters)
6) Pulmonary function test at day 28 and 90
   a. maximum vital capacity ($VC_{max}$)
   b. diffusing Capacity ($DL_{CO}$)
7) Difference in the change of mMRC (Modified Medical Research Council) Dyspnea Scale at day 28, day 90.
8) Changes of absolute lymphocyte counts and subsets, as well as cytokine/chemokine levels from baseline to day 6, 10, 28 and 90.

##### *3.4.3 Safety endpoints*

1) Adverse events
2) Serious adverse events
3) All-cause mortality

##### *3.4.4 Centralized imaging interpretation of endpoints based on high-resolution chest computed tomography imaging*

An independent central imaging reading and adjudication committee will be set for the evaluation of the four high-resolution chest CTs of every patient (baseline, day 10, 28, and 90). Centralized imaging interpretation process will be conducted separately when all patients finish 28-day follow-up (for baseline, day 10 and 28 chest CT) and 90-day follow-up (for day-90 chest CT). The reviewers will be blinded for the patient's visit information for the 28-day review and the treatment allocation for both reviews.

The review contains a software assisted lung volumetry and densitometry procedure and a manual review procedure.

**Software assisted lung volumetry and densitometry procedure**

1) Automatic lung segmentation: Import raw CT images to lung densitometry software. The software will automatically conduct lung segmentation for full lung area and lesion area.
2) Independent manual correction of segmentation: Two reviewers will manually correct the lesion segmentation by the software in itk-snap platform (V 3.8.0).
3) Volumetry and densitometry result output: if the difference in lesion proportion (%) of full lung volume between the two reviewers is no more than 10%, then the volumetry and densitometry will be recorded. The final result will be the average of the two reviewers. If the

difference is more than 10%, then a third reviewer with a higher level will be introduced in the review process. The third reviewer's result will be the final result.

**Lung volumetry and densitometry parameter:**

**Volumetry**

- Full lung volume (in $cm^3$, segmented and measured by software)
- Lesion volume (in $cm^3$, segmentation manually corrected by reviewers, measured by software)
- solid component lesion volume (in $cm^3$, segmented and measured by software)
- ground-glass lesion volume (in $cm^3$, segmented and measured by software)

**Densitometry** (measured by software)

- total voxel 'weight' in lesion area (segmentation manually corrected by reviewers, measured by software)

  voxel 'weight'=voxel density (in HU) × voxel volume (in voxel)
- volumes histogram of lung density distribution (<-750, -750~-300, -300~50, >50, measured by software)

**Manual review for pulmonary fibrosis – related morphological features**

The two independent reviewers will evaluate the following pulmonary fibrosis – related morphological features in CT scan at day 90. When results are different, the result of a third reviewer with a higher level will be the final call.

a. cord-like shadow
b. honeycomb-like shadows
c. interlobular septal thickening
d. intralobular interstitial thickening
e. pleural thickening

**Software**

- uAI-Discover-PNA, Version R001.0.0.15980, Shanghai united imaging intelligence Healthcare co. Ltd., China.

##### 3.5. Participant timeline

Figure. The schedule of enrolment, interventions, and assessments.

| STUDY PERIOD | Screening | Treatments | | | Follow-ups | | | |
|---|---|---|---|---|---|---|---|---|
| TIMEPOINT (DAY) | *-7~0* | 0 | 3 | 6 | *10* | *14** | *28#* | *90* |
| VISIT | *E* | *T1* | *T2* | *T3* | *F1* | *F2* | *F3* | *F4* |
| **ENROLMENT:** | | | | | | | | |
| **Informed consent** | X | | | | | | | |
| **Eligibility screen** | X | | | | | | | |
| ***Demographical characteristics*** | X | | | | | | | |
| ***Medical History*** | X | | | | | | | |
| **Urine pregnancy test (Women)** | X | | | | | | | X |
| **Urinalysis** | X | | | | | | | |
| **Stool analysis** | X | | | | | | | |
| **INTERVENTIONS:** | | | | | | | | |
| **Allocation** | | X | | | | | | |
| ***Experimental Group UC-MSCs*** | | X | X | X | | | | |
| ***Control Group UC-MSCs Placebo*** | | X | X | X | | | | |
| ***Both Groups Standard of Care*** | ⟶ | ⟶ | ⟶ | ⟶ | ⟶ | ⟶ | ⟶ | ⟶ |
| **ASSESSMENTS:** | | | | | | | | |
| ***Physical Examination*** | X | X | X | X | X | | X | X |
| ***Chest CT*** | X | | | | X | | X | X |
| ***mMRC Dyspnea Scale*** | | X | | X | X | | X | X |
| ***6-minute walk test*** | | | | | | | X | X |
| ***Pulmonary function (VC max and DLco)*** | | | | | | | X | X |

| | | | | | | | | |
|---|---|---|---|---|---|---|---|---|
| ***Blood oxygen saturation*** | X | | X | X | | | X | X |
| ***ECG*** | X | | | | | | X | X |
| ***Lab Tests***** | X | X | X | X | X | | X | X |
| ***Adverse event*** | → | → | → | → | → | → | → | → |
| ***Concomitant care*** | → | → | → | → | → | → | → | → |

* visit by phone call or phone app.
### Time window is from day28 to day35.
** including complete blood count, liver and kidney function, electrolyte panel, blood glucose, procalcitonin (PCT), Interleukin-6 (IL-6), creatine kinase (CK), troponin, myoglobin, brain natriuretic peptide(BNP), lactate dehydrogenase, D-dimer, C-Reactive Protein (CRP), absolute lymphocyte counts and subsets (including CD4 T, CD8 T, NK, B ), cytokine/chemokine levels (including IL-1β, IL-2, IL-4, IL-5, IL-6, IL-8, IL-10, IL-12P70, IL-17A, IL-17F, IL-22, TNF-α, TNF-β, IFN-γ, IL-1RA, IL-18, G-CSF, RANTES, MCP-1, IP-10, MIP-1α)

#### 3.6. Sample size

Due to the exploratory nature of this pilot trial and limited efficacy information of treatment in COVID-19 patients, no statistical hypothesis was made. The original target sample size was set to be 45 patients, with the allocation ratio of 2:1, 30 in the UC-MSCs group, and 15 in the placebo group. The enrolment soon reached the target number because of the rapid outbreak of COVID-19 in Wuhan city. Considering minimal serious adverse events were observed, we decided to expand the sample size to gathering more information from this study. The sample size first expanded to 60, then currently set at 90 with 60 patients in the UC-MSC group, 30 patients in the placebo group in the situation that fewer new COVID-19 patients were available since the outbreak was contained in Wuhan. Decisions were made in a manner to maintain the double-blind status of this study and approved by the institutional review board.

#### 3.7. Recruitment

Patient will be recruited from two designated hospitals for COVID-19 in Wuhan City, Hubei Province: Huoshenshan Hospital and Hubei Maternity and Child Health Care Hospital. No other recruitment strategy (e.g., advertisement) will be adopted in this study.

#### 3.8. Allocation, blinding and product management

To ensure the cytoactive of UC-MSCs, the stem cells need to be administrated to patients in 24 hours after production and require a strict storing condition (8-12 °C, light-sensitive). Moreover, UC-MSC packages are produced in Tianjin, which is more than 1000 kilometers away from Wuhan. To face the challenges in treatment allocation, product management, and logistics, a barcode tracking system will be introduced to this study. The system is developed upon a post-market medical product tracking system currently been used by national drug administration agencies, hospitals, and patients in China. The system will be embedded with drug tracking, management, shipping (including shipment position and storing temperature) based on unique barcodes of every bag and package. It will be connected to a web-based randomization system.

##### *3.8.1. Allocation and concealment of allocation*

The permutated-block randomization sequence is generated by a non-investigator of this study and uploaded to the randomization system. The patients will be randomly assigned to either the UC-MSC group or placebo group with a 2:1 allocation ratio stratified by sites. The concealment of the randomization sequence, block size will be ensured by the randomization system.

##### *3.8.2. Blinding*

This is a double-blind trial. Patients, investigators, and outcome assessors (independent central imaging reviewers) will all be blinded to treatment allocation. The blinding will be ensured by product making and packaging (the UC-MSC and the placebo cannot be identified by appearance and packaging). The unique barcode, package numbers are also non-informative. To further ensure the blinding procedure, each shipping package unit will include extra bags of UC-MSCs and placebos bags, and not all products will be used.

The emergency unblinding procedure will be triggered under the circumstances when knowledge of the patient's treatment allocation is necessary for handling an emergency (e.g., when a severe adverse event is observed, the investigator needs to know the treatment group for further management of the patients). When facing such circumstances, the investigator will log into the randomization system by web or by phone app and conduct the emergency unblinding procedure online. The system will record and notify the principal investigator for all unbinding activities.

##### *3.8.3. Product management*

In this trial, all study products (UC-MSC and placebo) logistics will be enhanced by the Product Identification Authentication and Tracking System (PIATS). The system is connected to the study's randomization system. When the enrollment process is ongoing, the PIATS will ensure the study site have enough product in-store. When storing is low, it will trigger a new batch of the products' shipping process.

The entire product management process: prepare, packaging, shipping, storing, and clinical administration to patients will be conducted and traced by barcode scanners. The barcode will be unique and also non-informative to the blinded personnel. Investigators can view production information, track shipping status, revive shipment, dispense to patients using the system, and the barcode scanner.

#### 3.9. Data collection and data management

This trial will be conducted under an infectious disease public health emergency, which makes clinical trial challenging in data collection and management. Non-healthcare personnel cannot be involved in getting direct contact with hospitalized patients with active COVID-19. Most of the work solely rely on doctors and nurses on-site, and they are already putting most of the effort on providing medical care for COVID-19 patients. There will not be additional assistance from a third party (e.g., contract research organizations) with data collection and on-site monitoring when patients are hospitalized, their work will only begin from the follow-up phase.

In this trial, most of the research data will be collected from electronic medical records (EMRs). Doctors and nurses in this study will follow a guidance to collect trial-necessary source data in combination with patients' routine medical records. All trial related EMRs will be authorized to export to an electronic source data platform to perform data integrating, standardizing and cleaning and verifying. Data derived from EMRs or non-EMR data will be manually collected by investigator (e.g. time to clinical improvement) or study coordinator.

The data of the centralized imaging interpretation will be directly exported from softwares in medical imaging workstation.

#### 3.10. Statistical consideration

##### 3.10.1. Statistical methods

This study is designed to be a phase II, exploratory clinical trial. We will focus on statistical description for statistical analyses. Because there are no pre-defined hypotheses made in this study, all statistical test, confidence intervals, P-value is for reference, not for inference.

Baseline characteristics will be provided using descriptive statistics as frequencies (percentage) or mean±SD.

The UC-MSCs group will be compared against the control. Both intention-to-treat (ITT) and per-protocol strategy will be adopted when analyzing primary and secondary outcomes. The full analysis set (FAS) will be considered as the primary analysis population. We will use chi-square test/fisher's exact test for binary outcomes, and t-test/Wilcoxon rank-sum test for continuous outcomes as appropriate. For comparison of changes from baseline between the two groups, an ANCOVA model considering baseline value as a covariate will be conducted in addition to direct comparison of the difference from baseline.

Adverse events, severer adverse events will be described in event frequency and proportion. Safety outcomes will be analyzed per treatment based on the safety population.

All statistical tests will be performed at a two-tailed test of $P<0.05$ statistically significant level. Analyses will be conducted using SAS software (version 9.4, NC, USA)

##### 3.10.2. Additional analyses

All detailed analyses will be pre-specified in the statistical analysis plan. Considering the exploratory nature of this trial, additional analyses may be applied. All additional analyses which not pre-specified will be noted as post-hoc analyses.

##### 3.10.3. Analysis population and missing data

Full Analysis Set (FAS): Including all randomized patients based on ITT principles. Only the following circumstances will be excluded: No UC-MSC or placebo are administrated after randomization.

Per-Protocol Set (PPS): Patients with major protocol violation that may affect the evaluation of primary endpoint.

Safety Set (SS): Patients with at least one dose of UC-MSC or placebo treatment

Baseline description will be performed in FAS, primary and secondary analysis will be performed in FAS and PPS population. Safety analysis will be performed in SS.

For primary analysis in FAS population, when the patient is missing a chest CT scan, the last scan's result will be carried to the missing visit. Other missing values of secondary outcomes and PP analyses will not be imputed.

#### 3.11. Safety/harms

Adverse events (AEs) and severer adverse events (SAEs) will be monitored and recorded from the time the subject signed informed consent to the completion of the follow-ups. AEs/SAEs will be recorded in detail, including onset date, duration, severity, treatment, relation to the investigational medical product. All AEs/SAEs will be followed up until finalized (recover/relief, stable, deaths, or other explainable circumstances, e.g., lost to follow-up).

SAEs, once identified, must be taken into action and reported within 24 hours. SAEs will be reported to IRB as soon as possible.

#### 3.12. Auditing

This study will be conducted following Good Clinical Practice (GCP). Investigators and coordinators will conduct source data verification when patients are hospitalized. Additional auditing and quality control by the site's GCP office and contract research organizations will be adopted. There will not be third party auditing in this trial during the COVID-19 outbreak, it will be available in the follow-up phase when all patients are discharged from hospital.

### 4. ETHICS AND DISSEMINATION

#### 4.1. Research ethics approval

This trial's protocol and informed consent forms have been approved by the institutional review boards of the Fifth Medical Center of PLA General Hospital. Major protocol amendments, SAEs suggestion will also be reported to IRB.

#### 4.2. Protocol amendments

Major changes, such as study objectives, study design, patient population, sample sizes, study procedures, outcomes which may impact potential benefit or harm of the patients will require a formal amendment to the protocol and will be approved by IRB.

#### 4.3. Informed consent process

The informed consent will be obtained from eligible hospitalized COVID-19 patients or their legal family substitute (for patients incapable of making an informed decision to give consent) by the investigators. Paper-based consent forms will be used in this study.

#### 4.4. Confidentiality

The investigators are bound to keep all patient's records that contain names or other personal identifiers confidential. All research data will be identified by a study subject ID only. Data or records shall not be used for purposes other than this clinical study.

#### 4.5. Declaration of interests

The principle investigators declare no financial and other competing interests.

#### 4.6. Access to data

All investigators from the steering committee will be given full access to the final data sets.

#### 4.7. Ancillary and post-trial care

The investigators will continue to follow up patients after 90 days for safety and research purposes. Long-period results of efficacy and safety will be collected.

#### 4.8. Dissemination policy

The study team will communicate trial results with health authorities, professionals, and patients who participated in this study. The results of this trial will be published when available.

Further publications, authorship of this study results must be reviewed by the principal investigator and sponsor, and written consent must be obtained.

Data sharing policy will be described in detail in the data sharing statement when the study result is published.

### 5. STUDY ADMINISTRATION

#### 5.1. Key contacts

**Central contact**

Principal Investigator: Fu-Sheng Wang, MD, Ph.D. 86-10-66933332

Investigator and coordinator: Lei Shi, MD, Ph.D. 86-10-66933333

Study Sites:
Optical Valley Branch of Maternal and Child Hospital of Hubei Province, Wuhan, Hubei, China, 430000
Contact: Wei-Fen Xie, MD, PhD

Wuhan Huoshenshan Hospital, Wuhan, Hubei, China, 430000
Contact: Lei Shi, MD, PhD

#### 5.2. Roles and responsibilities

##### *5.2.1. Protocol contributors*

Fu-Sheng Wang, Wei-Fen Xie, Chen Yao designed the trial.
Chen Yao developed randomization, data management, and statistical plan.
Lei Shi and Chongya Dong drafted the protocol.

##### *5.2.2. Sponsor and Funding and Collaborators*

**Sponsor**
The Fifth Medical Center of PLA General Hospital

**Funding**
1. National Key R&D Program of China (2020YFC0841900, 2020YFC0844000).
2. Innovation Groups of the National Natural Science Foundation of China (81721002).
3. National Science and Technology Major Project (2018ZX10302104-002).

**Collaborators**
Study sites:

Huoshenshan Hospital, Optical Valley Branch of Maternal and Child Hospital of Hubei Province, General Hospital of Central Theater Command (Follow-up only).

UC-MSC and Placebo Provider:
VCANBIO Cell & Gene Engineering Corp., Ltd, China.

Randomization system provider:
Chengdu Cims-medtech Co., Ltd, China.

Product Identification Authentication and Tracking System (PIATS) provider:
Alibaba Health Technology (China) Company Limited, China

Central imaging reading and adjudication provider:
Wuhan Union Hospital, Tongji Medical College of Huazhong University of Science and Technology, China

Electronic source data system and eCRF system provider:
Digital China Health Technologies Co., Ltd, China

Trial Monitoring/ Data management team/ Clinical Research Organization:
SciTrials Medical Technolody Ltd, China

Statistical analysis
Department of biostatistics, Peking University First Hospital, China

##### *5.2.3. Trial committees*

**Steering Committee:** Fu-Sheng Wang, Wei-Fen Xie, Chen Yao
**Operating Committee:** Xuechun Lu, Lei Shi, Hai Huang, Xiaojing Jiang, Liangliang Sun, Fanping Meng, Ming Shi, Lei Huang, Junliang Fu, Zhe Xu, Xin Yuan, Tianyi Liu, Chao Zhang, Wenjin Song, Yuanyuan Li, Ruonan Xu, Siyu Wang.
**Central Imaging Reading and Adjudication Committee**: Fan Yang, Lan Zhang, Ran Tao

### 7. APPENDICES

#### 7.1 Revision History

| Version | Date | Amendment Text | Description |
|---|---|---|---|
| 1.1 | March 6, 2020 | Sample size was expanded from 45 to 60. | The original target sample size was set to be 45 patients, with the allocation ratio of 2:1, 30 in the UC-MSCs group, and 15 in the placebo group. The enrolment soon reached the target number because of the rapid outbreak of COVID-19 in Wuhan city. Considering minimal serious adverse events were observed, we decided to expand the sample size to gathering more information from this study. The sample size first expanded to 60. |
| | | Single infusion of MSCs was altered to the same dose (4.0*10E7 cells per time). | The number of cells in a single infusion was changed from two different grades (body weight≥70Kg, 4.0*10E7 cells per time; body weight<70Kg, 3.0*10E7 cells per time ) to same grade. |
| 2.0 | March 11,2020 | Primary outcome measure was changed to size of lesion area and severity of pulmonary fibrosis by chest CT. | COVID-19 is an unprecedentedly new infectious disease in the world. The distribution and disease characteristics of patients are constantly developed. We adjusted the research protocol according to actual situation of enrolled patients. |
| | | Blood oxygen saturation, oxygenation index (PaO2/FiO2), CD4+ T cell count and cytokine level and side effects were added as secondary outcome measures. | |
| | | Improvement time of clinical critical treatment index within 28 days, all-cause mortality on Day 28, invasive mechanical ventilation rate and incidence of nosocomial infection were deleted in secondary outcome measures part. | |
| | | The following is added to the exclusion criteria:<br>• Interstitial lung damage caused by other reasons (in 2 weeks) | |

| | | | |
|---|---|---|---|
| | | • The pulmonary imaging revealed the interstitial damage of lungs before the COVID-19 confirmed. | |
| | | Maternal and Child Health Hospital of Hubei Province was added as collaborators. | |
| | | Sample size was expanded from 60 to 90. | |
| | | Age range was expanded to 75-years-old. | |
| 3.0 | March 23, 2020 | Official Title was changed to A Multicenter, Randomized, Double-blind, Placebo-controlled Study Evaluating the Efficacy and Safety of Human Mesenchymal Stem Cells in Combination With Standard Therapy in the Treatment of COVID-19 Patients With Severe Convalescence. | |
| | | General Hospital of Central Theater Command was added as collaborators. | The follow-up procedure will be conducted at the General Hospital of Central Theater Command of PLA. |
| | | mMRC (Modified Medical Research Council) dyspnea scale, 6-minute walk test, maximum vital capacity (VCmax), Diffusing Capacity (DLCO) were added as secondary outcome measures. | |
| | | Proportion of patients in each classification of clinical critical treatment index was deleted in secondary outcome measures part. | |
| 3.1 Final | June 1,2020 | Outcomes were confirmed as follows:<br>Imaging Endpoints by centralized interpretation<br>Primary endpoint measures (centralized imaging interpretation)<br>•Change in lesion proportion (%) of full lung volume from baseline to day 28. | |

| | | |
|---|---|---|
| | | Lesion proportion = lesion volume (in cm3) / full lung volume (in cm3)<br>Secondary endpoint measures (centralized imaging interpretation)<br>•Change in lesion proportion (%) of full lung volume from baseline to day 10 and 90<br>•Change in solid component lesion proportion (%) of full lung volume from baseline to day 10, 28 and 90.<br>•Change in ground-glass lesion proportion (%) of full lung volume from baseline to day 10, 28 and 90.<br>•Pulmonary fibrosis – related morphological features in CT scan at day 90<br>a. cord-like shadow<br>b. honeycomb-like shadows<br>c. interlobular septal thickening<br>d. intralobular interstitial thickening<br>e. pleural thickening<br>•Change in lung densitometry:<br>a.total voxel 'weight' in lesion area<br>voxel 'weight'=voxel density (in HU) × voxel volume (in voxel)<br>b. volumes histogram of lung density distribution (<-750, -750~-300, -300~50, >50)<br><br>3.4.2 Other secondary endpoints<br>1) Time to clinical recovery in 28 days.<br>Clinical improvement defined as a one-point deduction from baseline in a 6 ordinal scale:<br>1.Not hospitalized;<br>2.Hospitalized, not requiring supplemental oxygen;<br>3.Hospitalized, requiring supplemental oxygen;<br>4.Hospitalized, on non-invasive ventilation or high flow oxygen devices;<br>5.Hospitalized, on invasive mechanical ventilation or ECMO;<br>6.Death. |

| | | |
|---|---|---|
| | | 2)Change in oxygenation index (PaO2/FiO2) from baseline to day 6, 10, and 28.<br>3)The duration of oxygen therapy (in days)<br>4)Change in oxygen saturation from baseline to day 6, 10 and 28<br>5)Difference in 6-minute walk test at day 28 and day 90 (in meters)<br>6)Pulmonary function test at day 28 and day90<br>a. maximum vital capacity (VCmax)<br>b. diffusing Capacity (DLCO)<br>7)Difference in the change of mMRC (Modified Medical Research Council) Dyspnea Scale at day 28, day 90.<br>8)Changes of absolute lymphocyte counts and subsets, as well as cytokine/chemokine levels at baseline, day 6, day 10, day 28 and , day 90.<br><br>3.4.3 Safety endpoints<br>1) Adverse events<br>2) Serious adverse events<br>3) All-cause mortality |

#### *7.2 mMRC (Modified Medical Research Council) Dyspnea Scale*

Walking should be assessed on level ground

| | |
|---|---|
| Dyspnea only with strenuous exercise | 0 |
| Dyspnea when hurrying or walking up a slight hill | +1 |
| Walks slower than people of the same age because of dyspnea or has to stop for breath when walking at own pace | +2 |
| Stops for breath after walking 100 yards (91 m) or after a few minutes | +3 |
| Too dyspneic to leave house or breathless when dressing | +4 |
