## Supplement 2-SAP for "Treatment with human umbilical cord-derived mesenchymal stem cells for COVID-19 patients with lung damage: a randomised, double-blind, placebo-controlled phase 2 trial"

### A Randomized double-blind, placebo-controlled evaluation of the efficacy and safety of human mesenchymal stem cells combined with standard therapies in the treatment of patients with severe COVID-19

### Statistical Analysis Plan

Version number: v1.0 2020-06-05

Sponsor: The Fifth Medical Center of PLA General Hospital

Statistical unit: Peking University Clinical Research Institute

### Signature Page

**The authors:**

Yu Yongpei, statistician

| Signature | Date |
|---|---|

**Report reviewer, the person in charge of quality control:**

Yan Xiaoyan, Senior Statistician

| Signature | Date |
|---|---|

#### 1. Table of Abbreviations

| Abbreviations | Definition |
|---|---|
| AE | Adverse event |
| ALT | Alanine aminotransferase |
| AST | Aspartate aminotransferase |
| BMI | Body mass index |
| BUN | Blood urea nitrogen |
| CI | Confidence interval |
| CK | Creatine kinase |
| CMH | Cochran-Mantel-Haenszel test |
| Cr | Creatinine |
| CRC | Clinical research coordinator |
| CRF | Case report form |
| CRO | Contract research organization |
| CRP | C-reactive protein |
| CT | Computed tomography |
| CTA | Clinical trial agreement |
| DMC | Data Monitoring Committee |
| EC | Ethics committee |
| EMR | Electronic medical record |
| F | F statistic (result of covariance analysis) |
| FAS | Full analysis set |
| FiO2 | Fraction of inspiration O2 (oxygen absorption concentration) |
| GCP | Good clinical practices |
| ICF | Informed consent form |
| IL-6 | Interleukin-6 |
| ITT | Intent-to-treat analysis |
| IWRS | Interactive Web Response System (the central randomization system) |
| LOCF | Last observation carried forward |
| P | P value |
| PI | Principle investigator |
| PPS | Per-protocol set |
| RR | Breath rate |
| SAE | Serious adverse event |
| SD | Standard deviation |
| SOC | Standard of care |
| SOP | Standard operating procedures |
| SS | Safety data set |
| TEAE | Treatment emergent adverse event |
| UC-MSC | Umbilical cord mesenchymal stem cells |

#### 2. Research title

A randomized double-blind, placebo-controlled evaluation of the efficacy and safety of human mesenchymal stem cells combined with standard therapies in the treatment of patients with severe COVID-19

#### 3. Research objectives

To evaluate the efficacy and safety of umbilical cord-derived human mesenchymal stem cells in the treatment of severe COVID-19 patients.

#### 4. Trial design

##### 4.1 Study design

This study is a multi-center, prospective, randomized double-blind, placebo-controlled clinical trial. The overall study design is demonstrated in the figure below:

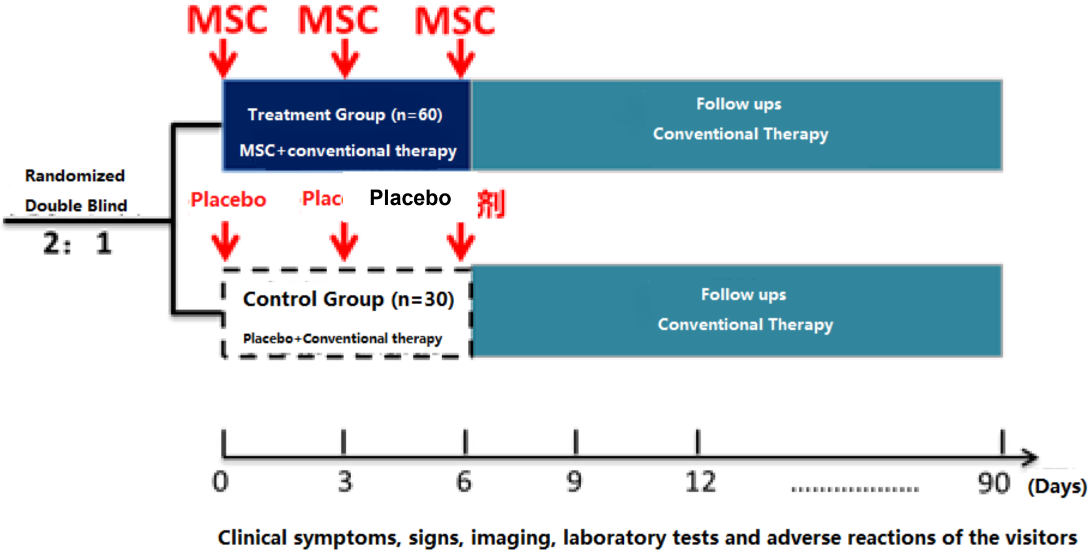

##### 4.2 Study intervention

The allocation ratio is 2:1 in the treatment group comparing to the control group.

The treatment group (60 cases): standard treatment + human umbilical cord mesenchymal stem cell (UC-MSC) treatment (intravenous injection once every 3 days for a total of 3 times (starting from day 0), the dose of each treatment is $4.0 \times 10^7$ MSCs with a volume of 100ml).

The control group (30 cases): standard treatment plan + placebo.

Standard treatment (SOC) will be implemented according to the "Chinese Clinical Guidance for COVID-19 Pneumonia Diagnosis and Treatment ($7^{th}$ edition)" compiled by the National Health Commission of China.

##### 4.3 Randomization and blinding

In order to reduce the possible selection bias and curative effect evaluation bias from investigator and subjects, this study will adopt a randomized double-blind, placebo-controlled design. Since all the patients would receive current standard care, the study has embedded patients' interests in the design.

This study intends to use the CIMS Interactive Web Response System (IWRS), which will be docked with the "Mashangfangxin" drug blinding system by Alibaba Health. Generally, in preventing the investigators, clinical teams or others relevant personnel from accessing the trial data, the Alibaba health system will remain inaccessible until the trial is completed. However, should circumstances arise, the Alibaba Health system also allows the emergency unblinding of individual subjects for the investigator.

To enable double-blinding, the medication of interest (the UC-MSC intravenous injections) must go through a scanning device that records the unique barcode on the medication package before use. Normally, unblinding is not allowed until all subjects have completed the study and the database is locked. Unblinding is allowed, however, when it is necessary to know the subject's treatment status to take special emergency treatment or actions. At this time, the investigator can log in to the IWRS to gather information on the subject in the emergent situation. The investigator is advised to contact the project team as much as possible before the unblinding and discuss the details. The phone of the project team or its designated personnel will be kept open 24 hours a day, 7 days a week. If the unblinding has been done, the project team must be notified as soon as possible. The date, time and reason for unblinding must be recorded in the relevant chapters and original documents of the IWRS, as well as on the electronic Case Report Form (eCRF). The unblinding document received from IWRS must be securely stored with the subject's original document.

#### 4.4 Research schedule

| **STUDY PERIOD** | **Screening** | **Treatments** | | | **Follow-ups** | | | |
|---|---|---|---|---|---|---|---|---|
| Time points | -7~0 days | 0 days | 3 days | 6 days | 10 days | 14~21 days | 28~35 days | 90 days |
| Candidate Enrollment Screening Form | √ | | | | | | | |
| Sign informed consent | √ | | | | | | | |
| Demographic data | √ | | | | | | | |
| Vital signs, physical examination | √ | √ | √ | √ | √ | | √ | √ |
| Past medical history | √ | | | | | | | |
| Infusion record | | √ | √ | √ | | | | |
| Blood routine | √ | | | √ | √ | | √ | √ |
| Procalcitonin | √ | | | √ | √ | | √ | √ |
| 6-minute walk test | | | | | | | √ | √ |
| IL-6 | √ | | | √ | √ | | √ | √ |
| Liver and kidney function, electrolytes, blood sugar | √ | | | √ | √ | | √ | √ |
| CK, LACTIC ACID | √ | | | √ | √ | | √ | √ |
| D DIMER | √ | | | √ | √ | | √ | √ |

| | | | | | | | | |
|---|---|---|---|---|---|---|---|---|
| Cardiac Function | √ | | | √ | √ | | √ | √ |
| CRP | √ | | | √ | √ | | √ | √ |
| ECG | √ | | | | | | √ | √ |
| Chest CT | √ | | | | √ | | √ | √ |
| mMRC dyspnea score | √ | √ | | √ | √ | | √ | √ |
| Forced vital capacity (VC max) | | | | | | | √ | √ |
| Lung diffusion function (DL co) | | | | | | | √ | √ |
| Lymphocyte subsets | √ | | | √ | √ | | √ | √ |
| Blood gas analysis | √ | | | √ | √ | | √ | √ |
| Urine routine | √ | | | | | | | |
| Stool routine | √ | | | | | | | |
| Adverse events and concomitant medication | √ | √ | √ | √ | √ | √ | √ | √ |
| Telephone follow-up | | | | | | √ | | |

#### 4.5 Inclusion criteria

##### 4.5.1 **Inclusion criteria**

(1) Age ≥18 and <75 years;

(2) Hospitalized;

(3) Severe COVID-19:

1) Laboratory confirmation of SARS-CoV-2 infection by reverse-transcription polymerase chain reaction (RT-PCR) from any diagnostic sampling source
2) Confirmed pneumonia by chest computed tomography imaging
3) Apply to any one of the following after onset: 1)dyspnea (RR ≥ 30 times/min), 2)finger oxygen saturation ≤ 93% in resting state, 3)arterial oxygen partial pressure (PaO2) / oxygen absorption concentration (FiO2) ≤ 300MMHG, 4)pulmonary imaging shows that the focus progress > 50% in 24-48 hours

(4) Confirmed interstitial lung damage by chest computed tomography imaging.

##### 4.5.2 **Exclusion criteria**

Subjects who meet any one of the following conditions will not be selected for the trial:

(1) Pregnancy or lactation;

(2) Patients with malignant tumor, other serious systemic diseases, or psychosis;

(3) Patients who are participating in other clinical trials;

(4) There was evidence of drug addiction within one year before joining the study;

(5) Unable or unwilling to sign the informed consent or comply with test requirements;

(6) Co-Infection of HIV, tuberculosis, influenza virus, adenovirus, or other respiratory infection viruses;

(7) Interstitial lung damage caused by other reasons (in 2 weeks);

(8) The pulmonary imaging revealed the interstitial damage of lungs before the COVID-19 confirmed;

(9) Mild, normal, or critical COVID-19 patients.

###### 4.5.3 **Withdrawal criteria**

Those who meet the following criteria will withdraw from the trial:

(1) Subjects withdraw informed consent, etc.;

(2) Poor compliance results in incomplete administration of UC-MSCs per protocol.

(3) During the treatment, if the subject experienced certain comorbidities and complications, AE and SAE or special physiological changes, the investigator may decide that it was not suitable to continue the treatment. Any grade 4 AE or laboratory abnormalities related to UC-MSCs treatment, it is up to the investigator to continue the treatment of any other AE, laboratory abnormalities or concomitant diseases that are not good to the subject;

(4) Any other event judged by the investigator that the patient should be withdrawn from the study.

If a subject withdraws from the study due to AE or abnormal laboratory test results, this important special event and the test results should be recorded in the medical record.

The reasons for the dropped cases should be recorded in detail, the safety and efficacy data of the subject should be obtained as much as possible, and all the end-of-study evaluations should be carried out with the consent and compliance of the subject. All case data should be kept complete for future reference.

###### 4.5.4 **Termination criteria**

The trial should be terminated if the following criteria are met:

(1) Serious safety problems occurred during the trial;

(2) The unit in charge of clinical research requests to terminate the trial (such as funding reasons, management reasons, etc.);

(3) The ethics committee or the administrative department request the termination of the trial;

(4) As the epidemic subsided, it is difficult to enroll new cases.

###### 4.5.5 **Complete research**

It is expected that 90 subjects will be enrolled. Subjects will be deemed to have completed the trial after completing all the treatments and all contents specified during the follow-up period (from the screening period to the last follow-up) according to the trial protocol.

###### 4.5.6 Withdraw from research

The subjects can withdraw from the study at any time during the study for any reason (special or non-special) without being unfairly treated. Subjects will not be restricted in their treatment and may continue to receive conventional treatment or other suitable treatment methods.

For subjects who are deemed ineligible in the screening period, the check items in the screening period should be stopped, and the reason for the screening failure should be recorded on the study completion/termination page. If the screened subjects withdraw from the trial, they should try to complete the check items in the current visit. And the reason for withdrawing from the study is recorded on the study completion/termination page after the study is completed.

#### 5. Data management

##### 5.1 Creating database

A database system project corresponding to the trial protocol will be created. The logical conditions are put in for the relevant items. The database preparation will be completed after the database passed the verification test.

##### 5.2 Data input

The staff responsible for the data input will first receive training on the use of the database system. The trained staff will synchronize the data entry. A data administrator will review the data entries and produce a query list for the potentially problematic entries. The data entry staff will then check the original data records to verify or revise the problematic entries. Data administrators will supervise the progress of data entry and verify the data regularly during the project to ensure data quality.

#### 6. Outcomes

##### 6.1 Primary endpoint measures

Primary endpoint measure: changes in the high-resolution CT imaging: changes in the size, location and nature of the lesions of the lung (such as strip-like, ground-glass, grid-like, solid component, etc.).

Imaging evaluation method: Four CT images (screening period, follow-up period of 10 days, 28-35 days, and 90 days) were evaluated by the independent imaging evaluation committee of the study center.

##### 6.2 Secondary endpoint measures

Secondary endpoints:

(1) Proportion of patients in each category on the 6-point scale at 7, 14, 28, and 90 days after randomization

(2) Blood gas analysis and oxygenation index;

(3) Duration of oxygen therapy (days);

(4) Finger pulse oxygen in resting state;

(5)6-minute walking experiment (before discharge);

(6) Lung function;

(7) mMRC dyspnea score;

(8) Features of immunological changes (changes in T lymphocytes and inflammatory factors, etc.);

#### 6.3 Safety endpoints

No formal hypothesis testing is performed for safety analysis. Descriptive statistics will be used to summarize safety events. Safety evaluation includes the following:

- Adverse events

Collect the frequency and nature of postoperative adverse events, and evaluate the safety.

### 7. Statistical Analysis

#### 7.1 Analysis data set

##### 7.1.1 Full Analysis Set (FAS)

On the basis of the ITT principle, all randomized subjects are included as much as possible. Only the following subjects who have been randomized are excluded: violation of important core inclusion criteria; subjects not receiving treatment with experimental drugs; there are no observed data after randomization.

The FAS population is the main group of efficacy evaluation in this study.

##### 7.1.2 Per Protocol Set (PPS)

On the basis of the FAS population, exclude subjects with protocol violations that may affect the efficacy evaluation.

The PPS population is the secondary population for the efficacy evaluation of this study.

##### 7.1.3 Safety Set (SS)

Including all patients who received at least 1 study drug treatment.

#### 7.2 Missing data processing

Last Observation Carry Forward (LOCF) is used for the absence of major efficacy indicators in the FAS population. The missing data on demographic information, secondary efficacy indicators, and safety indicators are not processed.

#### 7.3 Statistical Methods

##### 7.3.1 Principles of Statistical Analysis

The software used for statistical analysis is SAS 9.4 or above. The sample size calculation uses PASS13 software. The main and secondary outcomes are analyzed using the full analysis set and the per protocol set, and the safety data set is used to analyze safety indicators.

Unless otherwise specified, all significance tests and hypothesis tests use two-sided tests at the 5% significance level. Where appropriate, the obtained p-value will be quoted and a 95% two-sided confidence interval will be generated. All The p-value will retain three decimal places. In all tables, p-values less than 0.001

will be expressed as "<0.001".

Continuous variables will be summarized with summary statistics such as mean, standard deviation, median, minimum, maximum, lower quartile (Q1), upper quartile (Q3), etc. Categorical and ordinal variables will be summarized in terms of frequency and percentage.

The comparison between the two groups of general conditions will be analyzed by appropriate methods based on the type of indicators. The two-sample t test (if the data is normally distributed) or the Wilcoxon rank sum test (if the data is not normally distributed) will be used for the comparison of the two continuous indicators. Categorical data will be analyzed by the Chi-square test or exact probability method (if Chi-square test is not applicable); the grade data will be analyzed by the Wilcoxon rank sum test or the CMH test.

###### 7.3.2 Completion status and demographic analysis

The number of people in two groups in the data set, the distribution of cases in each center, and the dropout rate are statistically described. A detailed list of reasons for data incompleteness will be compiled. The analysis of the completion of the trial will be based on all randomized subjects.

Statistical description and comparison between groups of baseline demographic information (including but not limited to age, gender, ethnicity, medical history, and medication history) are performed. Summary statistics will be utilized according to the nature of the variable (continuous variable or categorical variable).

The demographic characteristics (age, height, vital signs, etc.), medical history, and medication history of the patients are statistically described, and the age, height, weight, etc. of the two groups are compared to measure the comparability of the two groups. When the characteristics are statistically different, the analysis of the primary efficacy indicators needs to consider to adjust the statistically different baseline factors.

The analysis of baseline demographic information and vital signs will be based on the FAS.

###### 7.3.3 Baseline analyses

The baseline evaluation was performed on the FAS and PPS sets. The baseline analysis used the observation value that is the most recent to the treatment start time during the screening period and the day 0 of the treatment period.

The baseline 6-point scale and the mMRC dyspnea score are statistically described as qualitative indicators, and the Wilcoxon rank sum test is used for comparison between groups.

Baseline blood gas analysis, finger pulse oxygen at rest (referring to oxygen saturation), lung function (forced vital capacity, lung diffusion function, etc.), lymphocyte subsets, IL-6, d-dimer and CRP Quantitative indicators are statistically described and compared between groups.

###### 7.3.4 **Primary efficacy meansures**

The primary efficacy evaluation is performed on the FAS and PPS.

The primary efficacy measure of this study is the changes in high-resolution CT imaging. The relative changes from baseline between the two groups at 28 days after treatment are statistically described as quantitative indicators, and the t-test or the Wilcoxon rank sum test is used for comparison between groups. At the same time, the covariance model, including center and baseline as covariables, is constructed. The least-square mean (LSmean), inter-group difference and 95% confidence interval of the two groups were calculated.

###### 7.3.5 **Secondary efficacy measures**

The secondary efficacy evaluation is performed on the FAS and PPS.

(1) Proportion of patients in each category of the 6-point scale

According to the classification index, the 6-point scale for each follow-up is statistically described, and the Wilcoxon rank test is used for comparison between groups.

(2) Blood gas analysis and oxygenation index

The blood gas analysis indicators of each follow-up are statistically described and compared as quantitative indicators between groups.

(3) Duration of oxygen therapy (days)

The duration of oxygen therapy is statistically described and compared as quantitative indicators between groups.

(4) Finger pulse oxygen at rest

The finger pulse oxygen is statistically described and compared as quantitative indicators between groups in each follow-up resting state.

(5) 6-minute walk experiment

The follow-up 6-minute walk experiment is statistically described and compared as quantitative indicators between groups.

(6) Lung function

The pulmonary function indicators of each follow-up are statistically described and compared as quantitative indicators between groups.

(7) mMRC dyspnea score

The 6-point scale for each follow-up is statistically described as qualitative indicators, and the Wilcoxon rank test was used for comparison between groups.

(8) Features of immunological changes (changes in T lymphocytes and inflammatory factors, etc.)

The immunological indicators such as changes in T lymphocytes and inflammatory factors at each follow-up and the changes from baseline are statistically described and compared as quantitative indicators between groups.

##### 7.3.6 Safety analysis

The safety evaluation includes adverse events, laboratory examinations, electrocardiogram examinations and combined medications. The safety evaluation is carried out on the SS set.

**1) Adverse events:**

Describe the total adverse events, adverse events that led to the dropout, adverse reactions, and serious adverse events in the two groups will be reported respectively. The number of various events and the number of people who are associated with the events will be counted. The percentage of the people suffered from the adverse events in each group will be compared using the Fisher exact probability method.

**2) Laboratory examination and ECG examination**

The laboratory tests of clinical significance and the ECG examination at each measurement time point before and after the treatment is statistically described.

#### 8. Explanation of statistical analysis results
