## Supplement 3-12 for "Treatment with human umbilical cord-derived mesenchymal stem cells for COVID-19 patients with lung damage: a randomised, double-blind, placebo-controlled phase 2 trial"

**Supplement 3. Presentative data of viable cell analysis of MSC product after its preparation in Tianjin and before intravenous transfusion in Wuhan.**

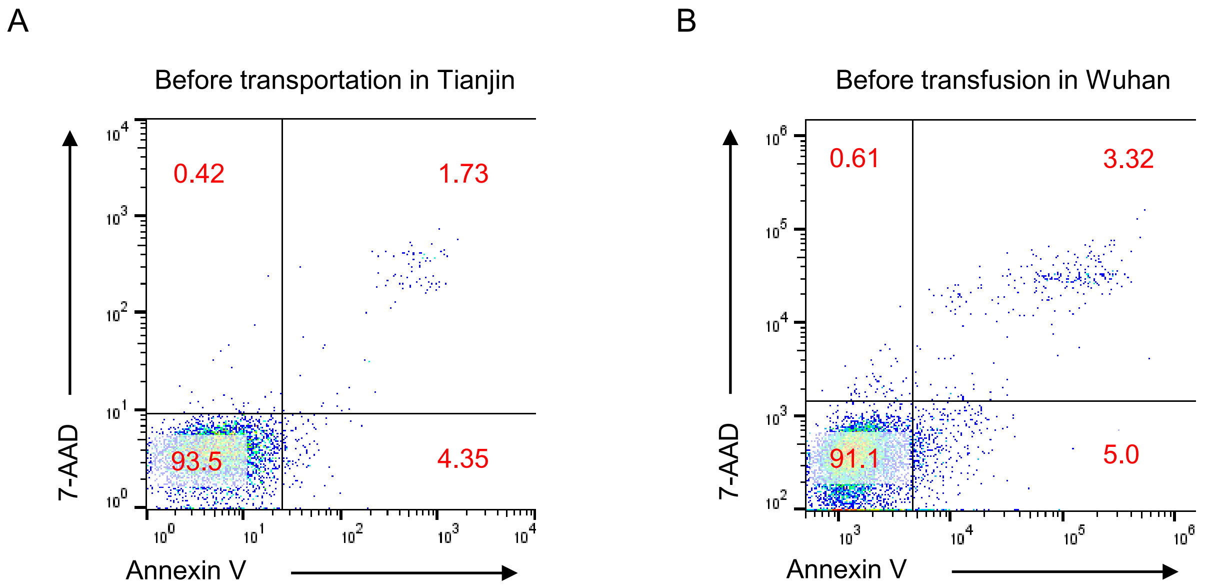


C

| Date | 0316 | 0318 | 0319 | 0320 | 0321 | 0322 | 0323 | 0324 | 0325 | 0326 |
|---|---|---|---|---|---|---|---|---|---|---|
| **Tianjin** | 93.9 | 95.0 | 93.9 | 95.3 | 94.7 | 94.7 | 93.3 | 96.8 | 89.6 | 96.3 |
| **Wuhan** | 87.8 | 87.9 | 79.8 | 92.2 | 90.6 | 93.1 | 84.9 | 93.4 | 80.9 | 93.4 |

(A) Representative flow cytometric plots for vitality test of UC-MSCs before transportation in Tianjin using 7-AAD/ Annexin V assay.

(B) Representative flow cytometric plots for vitality test of UC-MSCs before transfusion in Wuhan using 7-AAD/ Annexin V assay.

(C) Detailed results of cell vitalities after preparation in Tianjin and upon arrival in Wuhan.

**Supplement 4. Protocol deviation and per-protocol population.**

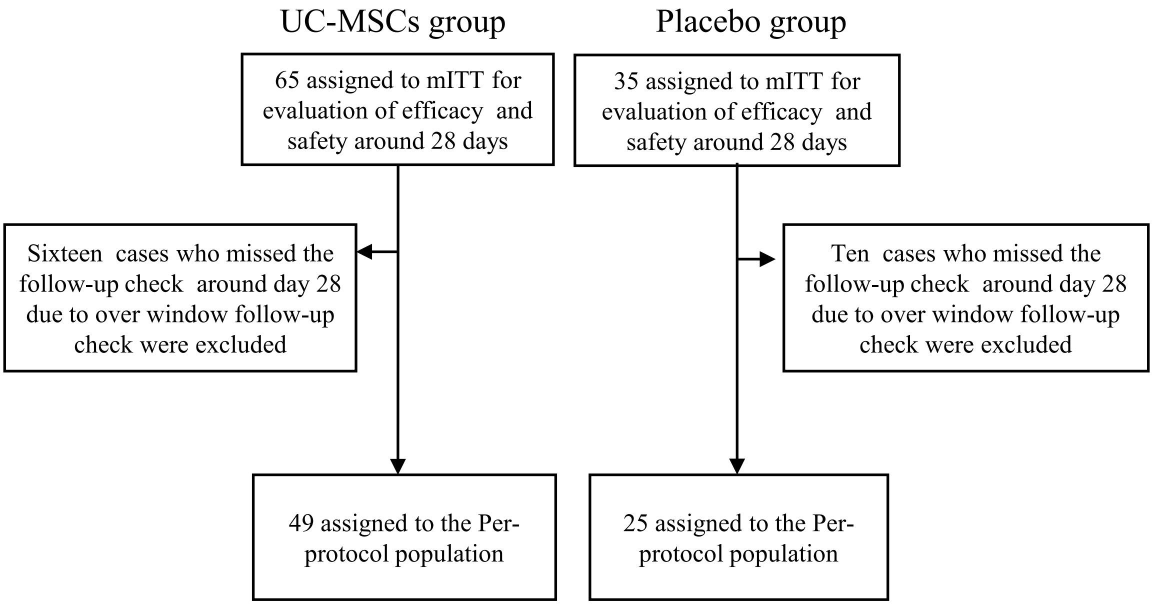

#### Supplement 5

### Table S1: Lesion proportion (%) of whole lung volume at day 0, 10, 28.

| | Day | UC-MSCs group (n=65) | Placebo group (n=35) | Difference |
|---|---|---|---|---|
| **Total lesion** proportion (%) of whole lung volume | 0 | 26.31(11.62,38.42) | 27.98(11.57,44.14) | -1.91(-10.66,5.38) † |
| | 10 | 25.99(6.32,39.95) | 25.24(11.62,47.12) | -2.24(-10.65,5.38) † |
| | 28 | 19.52(6.49,30.23) | 17.21(9.03,38.25) | -1.77(-9.50,5.31) † |
| **Solid component lesion** proportion (%) of whole lung volume | 0 | 2.59(0.69,5.20) | 2.52(0.77,4.91) | -0.05(-1.08,0.92) † |
| | 10 | 1.15(0.22,3.05) | 1.40(0.51,2.64) | -0.25(-0.86,0.27) † |
| | 28 | 0.94(0.22,2.25) | 1.45(0.51,2.66) | -0.32(-0.97,0.14) † |
| **Ground-glass lesion** proportion (%) of whole lung volume | 0 | 21.17(10.22,34.12) | 24.95(9.40,35.46) | -2.47(-9.82,4.37) † |
| | 10 | 23.01(6.11,36.90) | 23.22(10.21,40.22) | -2.22(-9.67,4.71) † |
| | 28 | 18.89(6.14,28.40) | 15.15(8.61,34.37) | -1.60(-8.60,5.40) † |

Data are expressed as median (interquartile range, IQR) The missing data has been imputed by LOCF(Last observation carried forward) strategy.

† Differences are expressed as Hodges-Lehmann estimator and 95% confidence interval (CI).

#### Supplement 6

#### Table S2: Baseline patient characteristics in the Per-protocol population.

| | UC-MSCs group (n=49) | Placebo group (n=25) |
|---|---|---|
| Age, years | 60.43(9.26) | 60.28(8.01) |
| Sex – no. (%) | | |
| Men | 27(55.10%) | 13(52.00%) |
| Women | 22(44.90%) | 12(48.00%) |
| Body Weight, kilogram* | 66.92(8.23) | 66.35(8.04) |
| BMI (Body Mass Index), Kg/m$^2$* | 24.89(3.14) | 25.11(3.11) |
| Time from symptom onset to starting study treatment, days | 45.00 (40.00,50.00) | 49.00 (45.00,55.00) |
| Any comorbidities | 25( 51.02%) | 13( 52.00%) |
| Hypertension | 13(26.53%) | 7(28.00%) |
| Diabetes | 9(18.37%) | 3(12.00%) |
| Chronic bronchitis | 2(4.08%) | 2(8.00%) |
| Chronic obstructive pulmonary disease | 2(4.08%) | 0(0.00%) |
| Concomitant medication | | |
| Antiviral drugs | 23(46.94%) | 15(60.00%) |
| Antibiotics | 18(36.73%) | 11(44.00%) |
| Glucocorticoid | 9(18.37%) | 6(24.00%) |
| Total lesion proportion (%): total lesion volume (in cm$^3$) / whole lung volume (in cm$^3$) | 24.82(11.41,41.76) | 27.28(12.16,45.32) |
| Solid component lesion proportion (%): Solid component lesion volume (in cm$^3$) / whole lung volume (in cm$^3$) | 2.02(0.61,5.49) | 2.20(0.76,4.61) |
| Six-category scale | | |
| 2-Hospitalized, not requiring supplemental oxygen | 11(22.45%) | 8(32.00%) |
| 3-Hospitalized, requiring supplemental oxygen | 37(75.51%) | 17(68.00%) |
| 4-Hospitalized, on noninvasive ventilation or high flow oxygen devices | 1(2.04%) | 0(0.00%) |
| White blood cell count ($10^9$/L) | 5.70(5.20,6.60) | 5.80(4.90,7.30) |

| | | |
|---|---|---|
| Lymphocyte count ($10^9$/L) | 1.39(1.19,1.80) | 1.47(1.24,1.84) |
| CD4 T (/μl) † | 643.00 (507.00,721.00) | 808.00 (465.00,1169.50) |
| CD8 T (/μl) † | 353.50 (275.00,484.00) | 411.00 (306.50,576.50) |
| B (/μl) † | 148.50 (112.00,215.00) | 182.50 (107.15,263.50) |
| NK (/μl) † | 198.00 (151.00,395.00) | 220.00 (133.00,337.50) |
| Neutrophil count ($10^9$/L) | 3.48(2.91,4.32) | 3.83(2.85,4.48) |
| Platelet count ($10^9$/L) | 214.00(174.00,255.00) | 210.00(176.00,247.00) |
| Hemoglobin （g/L） | 123.82(14.64) | 122.92(11.29) |
| D-dimer (mg/L) ‡ | 0.58(0.36,1.10) | 0.60(0.37,1.29) |
| IL-6 (pg/mL) § | 7.63 (6.08,9.78) | 8.76 (6.54,11.77) |
| CRP (mg/L) ¶ | 2.11 (1.16,3.47) | 1.38 (0.68,2.14) |
| SARS-CoV-2 test result‖ | | |
| SARS-Cov-2 IgG positive | 48(100.00%) | 24(100.00%) |
| SARS-Cov-2 IgM positive | 44( 91.67%) | 22( 91.67%) |
| SARS-Cov-2 nucleic acid detection positive | 36(73.47%) | 14(56.00%) |

Data are median (interquartile range (IQR)), n (%), ~~n/N(%),~~ or mean (SD)

* BMI values were available for 49 patients in the UC-MSCs group and 23 patients in the placebo group.

† CD4、CD8、CD19、CD56values were available for 46 patients in the UC-MSCs group and 24 patients in the placebo group.

‡ D-dimer values were available for 20 patients in the UC-MSCs group and 20 patients in the placebo group.

§ IL-6 values were available for 48 patients in the UC-MSCs group and 25 patients in the placebo group.

¶ CRP values were available for 21 patients in the UC-MSCs group and 10 patients in the placebo group.

‖ The test results are summarized from the hospitalization to the pre-random test. If there is any positive, it is defined as positive. . The IgG and IgM values were available for 48 patients in the UC-MSCs group and 24 patients in the placebo group.

#### Supplement 7

#### Table S3: Imaging outcomes in the per-protocol population.

| | **UC-MSCs group (n=49)** | **Placebo group (n=25)** | **Difference** |
|---|---|---|---|
| Change in **total** lesion proportion (%) of whole lung volume from baseline to day 28* | -19.32(-54.56,-3.74) | -8.13(-49.10,6.79) | -9.82(-29.26,9.41) |
| Change in **solid component** lesion proportion (%) of whole lung volume from baseline to day 28* | -54.29 (-73.44,-33.37) | -42.78 (-61.73,19.67) | -17.55(-41.04,1.88) |
| Change in **ground-glass** lesion proportion (%) of whole lung volume from baseline to day 28* | -18.46 (-57.32,5.61) | -6.30 (-45.69,8.05) | -6.39(-27.56,15.82) |

*Lesion proportion = lesion volume (in $cm^3$) / full lung volume (in $cm^3$). Data are median (interquartile range, IQR).

† Differences are expressed as Hodges-Lehmann estimator and 95% confidence interval (CI).

**Supplement 8. Dynamics of absolute counts of lymphocyte subsets and proinflammatory cytokines.**

A

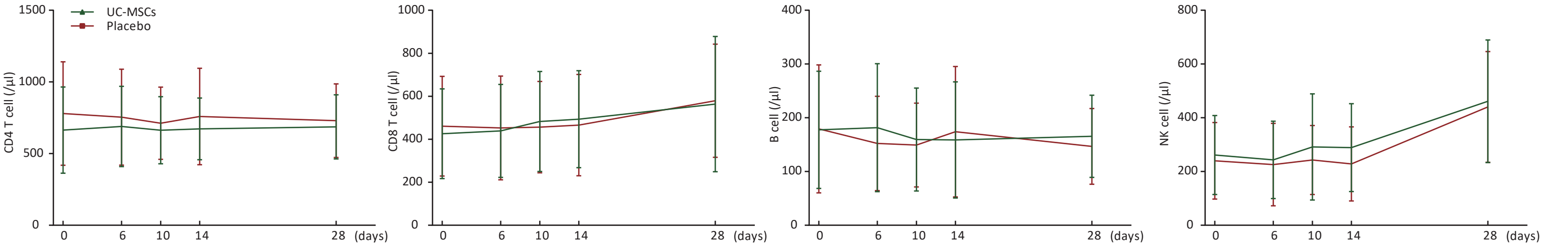


B

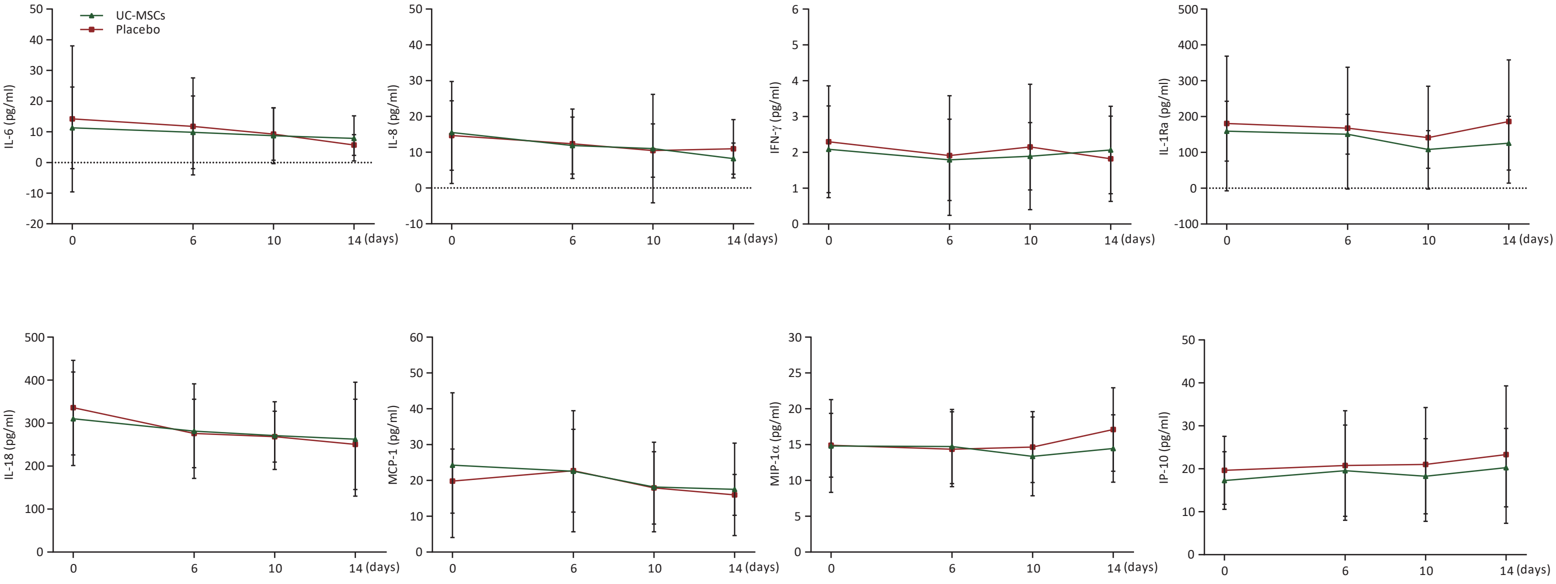


(A) Absolute counts of CD4 T, CD8 T, B and NK cells at day 0, 6, 10, and 14 after infusion.
(B) Plasma levels of IL-6, IL-8, IFN-γ, IL-1Ra, IL-18, MCP-1, MIP-1α, and IP-10 at day 0, 6, 10, and 14 after infusion.

**Supplement 9. Post Hoc Analyses for ensitivity analyses of primary end point.**

We have established five models in three analysis data sets (mITT, PPS and ITT) for sensitivity analyses of primary end point as follows. Refer to the following table 1-3 in details. The conclusions of all models were consistent with those of univariate analysis. Since this trial belonged to an exploratory study, the importance of statistical description was higher than that of statistical hypothesis test. Therefore, we did not add the results of multivariate model into the text of the manuscript.

Table 9-1. Analysis results of imaging outcomes for mITT based on the use of five models.
Table 9-2. Analysis results of imaging outcomes for PPS based on the use of five models.
Table 9-3. Analysis results of imaging outcomes for ITT based on the use of five models.

Table 9-1. Analysis results of imaging outcomes for mITT based on the use of five models.

| Model | UC-MSC group | Placebo group | Difference |
|---|---|---|---|
| Change in the total lesion proportion (%) of the whole lung volume from baseline to day 28 | | | |
| Model1 | -23.45(-33.24,-13.66) | -13.59(-26.93,-0.25) | -9.86(-26.41,6.68) |
| Model2 | -23.39(-33.23,-13.54) | -13.55(-26.95,-0.14) | -9.84(-26.46,6.78) |
| Model3 | -23.34(-33.22,-13.46) | -13.71(-27.18,-0.25) | -9.63(-26.33,7.07) |
| Model4 | -23.45(-33.24,-13.66) | -13.59(-26.93,-0.24) | -9.86(-26.41,6.68) |
| Model5 | -23.36(-33.19,-13.54) | -13.74(-27.13,-0.35) | -9.62(-26.24,6.99) |
| Change in solid component lesion proportion (%) of whole lung volume from baseline to day 28 | | | |
| Model1 | -47.69(-61.50,-33.89) | -21.83(-40.64,-3.03) | -25.86(-49.19,-2.53) |
| Model2 | -47.47(-61.30,-33.64) | -21.69(-40.53,-2.86) | -25.77(-49.14,-2.41) |
| Model3 | -47.47(-61.37,-33.56) | -21.70(-40.65,-2.75) | -25.77(-49.27,-2.26) |
| Model4 | -47.69(-61.50,-33.89) | -21.83(-40.64,-3.02) | -25.86(-49.19,-2.53) |
| Model5 | -47.65(-61.52,-33.77) | -21.92(-40.84,-3.00) | -25.73(-49.20,-2.26) |
| Change in ground-glass lesion proportion (%) of whole lung volume from baseline to day 28 | | | |
| Model1 | -14.80(-26.49,-3.10) | -7.73(-23.67,8.21) | -7.07(-26.84,12.70) |
| Model2 | -14.70(-26.46,-2.95) | -7.67(-23.68,8.34) | -7.03(-26.89,12.82) |
| Model3 | -14.68(-26.50,-2.87) | -7.74(-23.85,8.36) | -6.94(-26.92,13.04) |
| Model4 | -14.80(-26.49,-3.10) | -7.73(-23.67,8.21) | -7.07(-26.84,12.70) |
| Model5 | -14.75(-26.50,-2.99) | -7.82(-23.85,8.21) | -6.93(-26.81,12.96) |

The above analysis models are based on covariance model.

Treatment group factor was included in model 1, treatment group and center factors were included in model 2 as fixed effects, and treatment group, baseline and center factors were included as fixed effects in model 3. Center factor was included as random effect, treatment group factor was included as fixed effects in model 4, center factor was included as random effect, treatment group and baseline factors were included as fixed effects in model 5.

Table 9-2. Analysis results of imaging outcomes for PPS based on the use of five models.

| Model | UC-MSC group | Placebo group | Difference |
|---|---|---|---|
| Change in the total lesion proportion (%) of the whole lung volume from baseline to day 28 | | | |
| Model1 | -27.12(-37.82,-16.42) | -17.55(-32.68,-2.42) | -9.58(-28.11,8.96) |
| Model2 | -27.04(-37.81,-16.27) | -17.38(-32.62,-2.14) | -9.66(-28.31,8.99) |
| Model3 | -26.92(-37.66,-16.18) | -17.88(-33.09,-2.67) | -9.04(-27.66,9.58) |
| Model4 | -27.12(-37.83,-16.42) | -17.55(-32.68,-2.41) | -9.58(-28.11,8.96) |
| Model5 | -26.93(-37.59,-16.28) | -17.93(-33.00,-2.85) | -9.01(-27.48,9.46) |
| Change in solid component lesion proportion (%) of whole lung volume from baseline to day 28 | | | |
| Model1 | -44.28(-62.04,-26.52) | -10.09(-35.21,15.03) | -34.19(-64.96,-3.42) |
| Model2 | -44.02(-61.83,-26.21) | -9.57(-34.77,15.64) | -34.45(-65.29,-3.62) |
| Model3 | -43.97(-61.91,-26.03) | -9.77(-35.18,15.65) | -34.20(-65.30,-3.10) |
| Model4 | -44.28(-62.05,-26.51) | -10.09(-35.22,15.04) | -34.19(-64.96,-3.41) |
| Model5 | -44.15(-62.03,-26.28) | -10.35(-35.63,14.94) | -33.81(-64.78,-2.83) |
| Change in ground-glass lesion proportion (%) of whole lung volume from baseline to day 28 | | | |
| Model1 | -19.68(-32.18,-7.19) | -15.00(-32.67,2.67) | -4.68(-26.33,16.96) |
| Model2 | -19.56(-32.13,-6.99) | -14.76(-32.54,3.03) | -4.81(-26.57,16.96) |
| Model3 | -19.46(-32.06,-6.86) | -15.17(-33.02,2.68) | -4.29(-26.14,17.55) |
| Model4 | -19.68(-32.18,-7.18) | -15.00(-32.68,2.68) | -4.68(-26.34,16.97) |
| Model5 | -19.52(-32.03,-7.01) | -15.34(-33.04,2.36) | -4.18(-25.87,17.51) |

The above analysis models are based on covariance model.

Treatment group factor was included in model 1, treatment group and center factors were included in model 2 as fixed effects, and treatment group, baseline and center factors were included as fixed effects in model 3. Center factor was included as random effect, treatment group factor was included as fixed effects in model 4, center factor was included as random effect, treatment group and baseline factors were included as fixed effects in model 5.

Table 9-3. Analysis results of imaging outcomes for ITT based on the use of five models.

| Model | UC-MSC group | Placebo group | Difference |
|---|---|---|---|
| Change in the total lesion proportion (%) of the whole lung volume from baseline to day 28 | | | |
| Model1 | -23.09(-32.78,-13.41) | -13.59(-26.88,-0.29) | -9.51(-25.95,6.94) |
| Model2 | -23.05(-32.77,-13.32) | -13.54(-26.89,-0.19) | -9.51(-26.02,7.01) |
| Model3 | -22.99(-32.75,-13.23) | -13.70(-27.11,-0.29) | -9.29(-25.88,7.30) |
| Model4 | -23.09(-32.78,-13.41) | -13.59(-26.88,-0.29) | -9.51(-25.96,6.94) |
| Model5 | -23.01(-32.72,-13.30) | -13.74(-27.09,-0.40) | -9.27(-25.78,7.24) |
| Change in solid component lesion proportion (%) of whole lung volume from baseline to day 28 | | | |
| Model1 | -46.97(-60.65,-33.30) | -21.83(-40.61,-3.06) | -25.14(-48.37,-1.91) |
| Model2 | -46.81(-60.50,-33.12) | -21.68(-40.47,-2.89) | -25.13(-48.37,-1.88) |
| Model3 | -46.81(-60.57,-33.05) | -21.68(-40.59,-2.77) | -25.13(-48.52,-1.74) |
| Model4 | -46.97(-60.65,-33.29) | -21.83(-40.62,-3.05) | -25.14(-48.37,-1.90) |
| Model5 | -46.93(-60.67,-33.18) | -21.92(-40.81,-3.04) | -25.00(-48.37,-1.64) |
| Change in ground-glass lesion proportion (%) of whole lung volume from baseline to day 28 | | | |
| Model1 | -14.57(-26.12,-3.02) | -7.73(-23.59,8.14) | -6.84(-26.47,12.78) |
| Model2 | -14.51(-26.11,-2.90) | -7.67(-23.60,8.27) | -6.84(-26.55,12.87) |
| Model3 | -14.48(-26.15,-2.82) | -7.74(-23.76,8.29) | -6.75(-26.57,13.08) |
| Model4 | -14.57(-26.13,-3.02) | -7.73(-23.59,8.14) | -6.84(-26.47,12.78) |
| Model5 | -14.52(-26.14,-2.91) | -7.82(-23.77,8.13) | -6.70(-26.44,13.03) |

The above analysis models are based on covariance model.

Treatment group factor was included in model 1, treatment group and center factors were included in model 2 as fixed effects, and treatment group, baseline and center factors were included as fixed effects in model 3. Center factor was included as random effect, treatment group factor was included as fixed effects in model 4, center factor was included as random effect, treatment group and baseline factors were included as fixed effects in model 5.

#### Supplement 10

##### Six-category scale for COVID-19 patients

| Six-category scale for COVID-19 patients | |
|---|---|
| 1 | discharge (alive) |
| 2 | hospital admission, not requiring supplemental oxygen |
| 3 | hospital admission, requiring supplemental oxygen |
| 4 | hospital admission, requiring high-flow nasal cannula or non-invasive mechanical ventilation |
| 5 | hospital admission, requiring extracorporeal membrane oxygenation or invasive mechanical ventilation |
| 6 | death |

#### Supplement 11

### Normal Range of Lab Test

| | Normal range |
|---|---|
| White blood cell count ($10^9$/L) | 3.5-9.5 |
| Lymphocyte count ($10^9$/L) | 1.1-3.2 |
| CD4 T cells (/µl) | 550-1440 |
| CD8 T cells (/µl) | 320-1250 |
| B cells (/µl) | 90-560 |
| NK cells (/µl) | 150-1100 |
| Neutrophil count ($10^9$/L) | 1.8-6.3 |
| Platelet count ($10^9$/L) | 125-350 |
| Haemoglobin (g/L） | 316-354 |
| D-dimer (mg/L) | 0-0.55 |
| CRP (mg/L) | 0-4 |
| IL-6 (pg/ml) | 0-11.09 |
| IL-8 (pg/ml) | 0-15.71 |
| IFN-γ (pg/ml) | 0-4.43 |
| IL-1Ra (pg/ml) | 131.6-340.01 |
| IL-18 (pg/ml) | 57.8-364.4 |
| MCP-1 (pg/ml) | 0-36.66 |
| MIP-1α (pg/ml) | 0-18.99 |
| IP-10 (pg/ml) | 0-23.47 |

Print Form



**Table.** CONSORT 2010 Checklist of Information to Include When Reporting a Randomized Trial[a]

| Section and Topic | Item No. | Checklist Item | Reported on Page No. |
|---|---|---|---|
| **Title and abstract** | | | |
| | 1a | Identification as a randomized trial in the title | 1 |
| | 1b | Structured summary of trial design, methods, results, and conclusions (for specific guidance see CONSORT for abstracts) | 4-5 |
| **Introduction** | | | |
| Background and objectives | 2a | Scientific background and explanation of rationale | 7 |
| | 2b | Specific objectives or hypotheses | 7 |
| **Methods** | | | |
| Trial design | 3a | Description of trial design (such as parallel, factorial) including allocation ratio | 8-9 |
| | 3b | Important changes to methods after trial commencement (such as eligibility criteria), with reasons | NA |
| Participants | 4a | Eligibility criteria for participants | 8 |
| | 4b | Settings and locations where the data were collected | 8 |
| Interventions | 5 | The interventions for each group with sufficient details to allow replication, including how and when they were actually administered | 9-11 |
| Outcomes | 6a | Completely defined prespecified primary and secondary outcome measures, including how and when they were assessed | 11 |
| | 6b | Any changes to trial outcomes after the trial commenced, with reasons | Supplem |
| Sample size | 7a | How sample size was determined | 12 |
| | 7b | When applicable, explanation of any interim analyses and stopping guidelines | NA |
| Randomization | | | |
| Sequence generation | 8a | Method used to generate the random allocation sequence | 9,Supple |
| | 8b | Type of randomization; details of any restriction (such as blocking and block size) | 9,Supple |
| Allocation concealment mechanism | 9 | Mechanism used to implement the random allocation sequence (such as sequentially numbered containers), describing any steps taken to conceal the sequence until interventions were assigned | 9,Supple |
| Implementation | 10 | Who generated the random allocation sequence, who enrolled participants, and who assigned participants to interventions | 9,Supple |
| Blinding | 11a | If done, who was blinded after assignment to interventions (for example, participants, care providers, those assessing outcomes) and how | 9,Supple |
| | 11b | If relevant, description of the similarity of interventions | 9,Supple |
| Statistical methods | 12a | Statistical methods used to compare groups for primary and secondary outcomes | 12 |
| | 12b | Methods for additional analyses, such as subgroup analyses and adjusted analyses | 12 |
| **Results** | | | |
| Participant flow (a diagram is strongly recommended) | 13a | For each group, the numbers of participants who were randomly assigned, received intended treatment, and were analyzed for the primary outcome | 13,Figur |
| | 13b | For each group, losses and exclusions after randomization, together with reasons | 13 |
| Recruitment | 14a | Dates defining the periods of recruitment and follow-up | 13 |
| | 14b | Why the trial ended or was stopped | NA |
| Baseline data | 15 | A table showing baseline demographic and clinical characteristics for each group | 13-14, T |
| Numbers analyzed | 16 | For each group, number of participants (denominator) included in each analysis and whether the analysis was by original assigned groups | 13-14, F |
| Outcomes and estimation | 17a | For each primary and secondary outcome, results for each group, and the estimated effect size and its precision (such as 95% confidence interval) | 14-15 |
| | 17b | For binary outcomes, presentation of both absolute and relative effect sizes is recommended | NA |
| Ancillary analyses | 18 | Results of any other analyses performed, including subgroup analyses and adjusted analyses, distinguishing prespecified from exploratory | 14-15,Fi |
| Harms | 19 | All important harms or unintended effects in each group (for specific guidance see CONSORT for harms) | 15-16 |
| **Comment** | | | |
| Limitations | 20 | Trial limitations, addressing sources of potential bias, imprecision, and, if relevant, multiplicity of analyses | 18-19 |
| Generalizability | 21 | Generalizability (external validity, applicability) of the trial findings | 19 |
| Interpretation | 22 | Interpretation consistent with results, balancing benefits and harms, and considering other relevant evidence | 16-18 |
| **Other information** | | | |
| Registration | 23 | Registration number and name of trial registry | 5 |
| Protocol | 24 | Where the full trial protocol can be accessed, if available | Supplem |
| Funding | 25 | Sources of funding and other support (such as supply of drugs), role of funders | 20 |

[a] We strongly recommend reading this statement in conjunction with the CONSORT 2010 Explanation and Elaboration for important clarifications on all the items. If relevant, we also recommend reading CONSORT extensions for cluster randomized trials, noninferiority and equivalence trials, nonpharmacological treatments, herbal interventions, and pragmatic trials. Additional extensions are forthcoming: for those and for up-to-date references relevant to this checklist, see http://www.consort-statement.org.

©2010 American Medical Association. All rights reserved.
